# Using Large Language Models to Annotate Complex Cases of Social Determinants of Health in Longitudinal Clinical Records

**DOI:** 10.1101/2024.04.25.24306380

**Authors:** Alexandra Ralevski, Nadaa Taiyab, Michael Nossal, Lindsay Mico, Samantha N. Piekos, Jennifer Hadlock

## Abstract

Social Determinants of Health (SDoH) are an important part of the exposome and are known to have a large impact on variation in health outcomes. In particular, housing stability is known to be intricately linked to a patient’s health status, and pregnant women experiencing housing instability (HI) are known to have worse health outcomes. Most SDoH information is stored in electronic health records (EHRs) as free text (unstructured) clinical notes, which traditionally required natural language processing (NLP) for automatic identification of relevant text or keywords. A patient’s housing status can be ambiguous or subjective, and can change from note to note or within the same note, making it difficult to use existing NLP solutions. New developments in NLP allow researchers to prompt LLMs to perform complex, subjective annotation tasks that require reasoning that previously could only be attempted by human annotators. For example, large language models (LLMs) such as GPT (Generative Pre-trained Transformer) enable researchers to analyze complex, unstructured data using simple prompts. We used a secure platform within a large healthcare system to compare the ability of GPT-3.5 and GPT-4 to identify instances of both current and past housing instability, as well as general housing status, from 25,217 notes from 795 pregnant women. Results from these LLMs were compared with results from manual annotation, a named entity recognition (NER) model, and regular expressions (RegEx). We developed a chain-of-thought prompt requiring evidence and justification for each note from the LLMs, to help maximize the chances of finding relevant text related to HI while minimizing hallucinations and false positives. Compared with GPT-3.5 and the NER model, GPT-4 had the highest performance and had a much higher recall (0.924) than human annotators (0.702) in identifying patients experiencing current or past housing instability, although precision was lower (0.850) compared with human annotators (0.971). In most cases, the evidence output by GPT-4 was similar or identical to that of human annotators, and there was no evidence of hallucinations in any of the outputs from GPT-4. Most cases where the annotators and GPT-4 differed were ambiguous or subjective, such as “living in an apartment with too many people”. We also looked at GPT-4 performance on de-identified versions of the same notes and found that precision improved slightly (0.936 original, 0.939 de-identified), while recall dropped (0.781 original, 0.704 de-identified). This work demonstrates that, while manual annotation is likely to yield slightly more accurate results overall, LLMs, when compared with manual annotation, provide a scalable, cost-effective solution with the advantage of greater recall. At the same time, further evaluation is needed to address the risk of missed cases and bias in the initial selection of housing-related notes. Additionally, while it was possible to reduce confabulation, signs of unusual justifications remained. Given these factors, together with changes in both LLMs and charting over time, this approach is not yet appropriate for use as a fully-automated process. However, these results demonstrate the potential for using LLMs for computer-assisted annotation with human review, reducing cost and increasing recall. More efficient methods for obtaining structured SDoH data can help accelerate inclusion of exposome variables in biomedical research, and support healthcare systems in identifying patients who could benefit from proactive outreach.

## Introduction

The overwhelming majority of patients in the US have their data stored in electronic health records (EHRs). Information regarding a patient’s exposure to social determinants of health (SDoH), such as housing status, employment status, education, and quality of domestic life, provides relevant information that informs patient care and provides valuable avenues for intervention and treatment^1^. It has been estimated that SDoH can affect almost 50% of country-level variation in health outcomes, while clinical care impacts as little as 20%^2^. Housing data in particular, including a patient’s recent housing status, is known to be intricately linked to their health status^3–5^. Therefore, gaining insight into a patient’s current and past living situation is essential to providing more complete and equitable care. It is also important for research, where capturing longitudinal exposome data is essential for analysis of health outcomes.

Housing stability is known to exist on a continuum, from complete stability (access to housing of reasonable quality in the absence of threats) to complete instability (no access to housing of reasonable quality)^6^. It is well known that people who are experiencing housing instability (HI) are at greater risk for other health issues, including substance use, comorbidities, and mental illness^3,7,8^. People facing HI are also at an increased risk for homelessness^5,9,10^, which is associated with increased risk of morbidity and mortality^7^. Patients experiencing homelessness are also more likely to end up in the emergency department, have longer hospital stays than low-income housed persons, and are less likely to use preventive services^3,10^. Women who are experiencing HI while pregnant face additional challenges, as they usually require consistent access to care throughout their pregnancy. Adverse exposures prior to and during pregnancy can put a child at increased risk of both short- and long-term health consequences, and it is known that women who experience housing instability during pregnancy are at higher risk of adverse pregnancy outcomes, including preeclampsia, preterm birth, neonatal intensive care unit admission, and maternal morbidity^11–14^.

SDoH are rarely well documented in structured EHR data^15–17^. This leads to access barriers for researchers and caregivers. In addition, manually identifying SDoH, for example through chart abstraction, can be time-consuming and expensive, and infeasible to do at scale. Because structured data has often been optimized for purposes other than individual care or research, free-text descriptions capture greater breadth and complexity of a patient’s social and behavioral history. Existing projects, such as PRAPARE^18^ and emerging national interoperability plans^19^ are providing paths for clinicians to better capture SDoH data in structured fields, but widespread data standards for data harmonization are still in early development^20^.

Traditional NLP extraction of SDoH information from free-text notes has relied heavily on identification of keywords or phrases, using either manual or semi-automated lexicon curation, rule-based methods, or word embeddings^21–24^. However, most of these models are vulnerable to false positives and can only capture simplified concepts related to SDoH. In addition, previous research to identify housing instability from the EHR has focused primarily on homelessness or simplified housing-related concepts ^24–26^. However, because HI is heterogeneous with many intersecting dimensions, classifying a patient experiencing housing instability can be more complex than some aspects of social history and exposures, such as smoking. By contrast, large language models (LLMs) such as OpenAI’s GPT (Generative Pre-trained Transformer) models can handle large quantities of complex, unstructured data using only simple prompts. Research using LLMs on EHR data is still in its early stages, and most work has focused on either fine-tuning models for medical relevance^27,28^, comparing model performance to identify the presence or absence of SDoH statements^29^, or using LLMs for disease diagnosis or phenotyping^30,31^. In addition, it has not been made clear whether the quality of note text flagged as relevant by GPT is similar to that of a human annotator, and whether or not it is likely to contain hallucinatory text. LLMs such as GPT could also perpetuate health inequity if they perform differently for different patient populations. It is therefore important to test for bias in these models to inform future decisions regarding the use of LLMs in the healthcare setting. In addition, the possibility of using de-identified clinical notes for abstraction is an appealing for further supporting patient privacy. However, de-identification processes involve obfuscation of important details, including dates and locations. This may alter the semantic underpinnings of a given text, making it difficult for a LLM to accurately identify and label SDoH within a given note.

We examined whether LLMs were able to identify housing instability in clinical free-text notes with greater accuracy compared to manual annotation, regular expressions (RegEx), and a pre-trained named-entity recognition (NER) model for SDoH, using electronic health records for a population of pregnant women. We also examined the possibility of algorithmic bias in the predictions made by GPT-4 and GPT-3.5, as well as the differences in LLM performance on de-identified versions of patient notes. These methods have the potential to provide greater access to existing SDoH data that is valuable for retrospective research, chart review for prospective trials, and population health interventions to identify those who might benefit from proactive outreach.

## Methods

This retrospective study protocol was performed in compliance with the Health Insurance Portability and Accountability Act (HIPAA) Privacy Rule and was approved by the Institutional Review Board (IRB) at PSJH with Study Number 2020000783. Consent was waived because disclosure of protected health information for the study was determined to involve no more than a minimal risk to the privacy of individuals.

### Study setting and participants

Providence St Joseph Health (PSJH) is an integrated U.S. community healthcare system that provides care in urban and rural settings across seven states: Alaska, California, Montana, Oregon, New Mexico, Texas, and Washington. Using the PSJH electronic health records, we identified deliveries from June 8, 2010, through May 29, 2023 (n=595,600). We included singleton deliveries in a cohort of pregnant people aged 18-44 at the start of pregnancy (n=557,406) as previously described^32^. We limited the deliveries to records that had associated gravida, term, preterm, abortion, and living data (GTPAL) information, and to patients who received care in the PSJH system during pregnancy. For patients with more than one pregnancy episode, we randomly selected a single episode (n=408,158). We limited our patient cohort to those with complete Social Vulnerability Index (n=372,208) information as previously described^32^. To identify patients from our cohort who were experiencing housing instability (“preliminary positive class”), we searched for patients who had either a SNOMED code for housing instability (Supplementary Table S1) or a matching string for the word “homeless” in one or more of their free-text notes (n=13,024). Patients who did not meet these criteria were considered in the “preliminary negative class” (n=359,184) (Fig. 1).

**Fig. 1:**
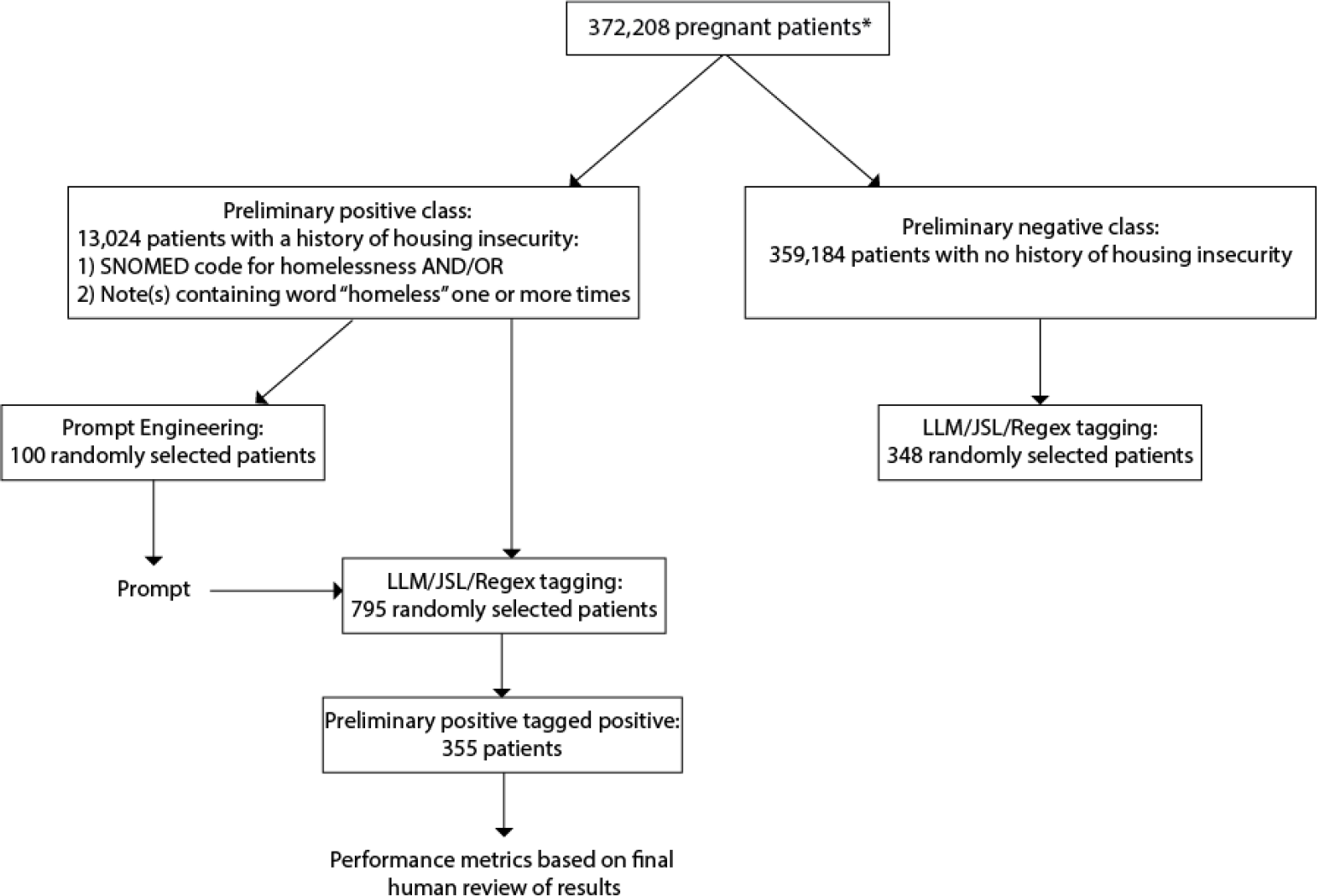
Cohort selection and experimental design. *Pregnant patients were selected as previously described^32^. Patients used for prompt engineering (100) and patients used for tagging (795) did not overlap.

### Task definition and data labeling

We defined different instances of housing stability and instability by first carrying out interviews with various subject matter experts (SMEs), including clinicians, social workers, and resource specialists. We generated preliminary annotation guidelines which were then iteratively refined and finalized with additional input from SMEs. The final annotation guidelines can be found in the Appendix. These guidelines distinguish between stable and unstable housing versus an *unknown* housing status, with examples for each pulled from EHR notes. An explicit definition of *history of housing instability* was also created. If a note contained any information on housing it had to be labeled as either *stably housed, current housing instability*, or *history of/past housing instability*. If the note contained no information on housing, it was labeled as *unknown*. These guidelines were then used to create the prompt used by the LLM. The annotation was divided into two rounds, with each reviewer annotating their own set of notes for the first round (*original label*). For the second round, 25% of the notes underwent dual annotation. If there was any disagreement for a given note between two annotators, a third annotator was asked to make a final decision. All the notes were then manually compared to results from GPT-4 to identify any notes that were obviously missed by reviewers. After the second round, a *final* label was assigned to each note. Performance metrics for manual annotation were calculated by comparing the *original* and *final* labels for each note. After manual annotation from reviewers, interrater reliability was calculated using Cohen’s Kappa on the notes that underwent dual annotation during the second round of annotation. The average time spent manually annotating a single note was calculated by each reviewer timing themselves for the time spent to annotate ten notes, and then taking the average. The two averages for each reviewer were then averaged.

### Models

Data processing was accomplished using the Azure AI Services API’s for GPT-4 and GPT-3.5 within the secure Providence cloud environment. GPT-4 version 0613 had a 32K token window, while GPT-3.5 Turbo version 0613 had a 16K token window. Both GPT models were run using LangChain and OpenAI libraries. The John Snow Labs (JSL) NER model (ner_sdoh_en_4.4.3_3.0_1686654976160, https://nlp.johnsnowlabs.com/2023/06/13/ner_sdoh_en.html) is a state-of-the-art SDoH model designed to detect and label SDoH entities within text data. The housing-specific label includes entities related to the conditions of the patient’s living spaces, for example: homeless, housing, small apartment, etc. JSL was run using sparknlp_jsl.version 5.0.0. To determine whether a RegEx search would identify relevant patient notes related to housing, we generated a preliminary list of keywords and phrases related to housing instability, which was then reviewed by SMEs to generate a final list (Supplementary Table S2).

### Prompt engineering

Our prompt was developed using chain-of-thought (CoT) prompting, where the problem/question description is initially stated and the LLM is asked to identify relevant evidence first, and then provide an answer. This method has been shown to be more accurate than asking the LLM to only provide an answer^33^. GPT-4 and GPT-3.5 were asked to first identify chunks of evidence verbatim from the text (*evidence*). The model was then asked to go through each of the four labels: *housing noted, housing instability current, housing stability current* and *housing instability history* and provide an answer for each. The model was then asked to provide a *justification* explaining why it chose a specific label and the LLMs were explicitly asked not to make up any information. GPT-3.5 was not used in the prompt engineering phase, and the prompt developed for GPT-4 was also used for GPT-3.5. The final prompt can be found in the Appendix.

For prompt engineering, we randomly selected 100 patients from the preliminary positive class and extracted all patient notes within one year of a patient’s conception date. Of the 100 patients, 51 of them had at least one note within one year of conception date, for a total of 1,569 notes (Fig. 1). Of those 51 patients, we used the JSL model to identify 70 notes from 16 patients related to housing. The 70 notes were then manually annotated by two independent researchers and the answers were compared. Any disagreement between the annotators was discussed and a final decision was made. From our annotation guidelines we developed an initial prompt for GPT-4 that contained definitions of housing instability as stated in the annotation guidelines, as well as examples of housing instability from samples found in the patient notes. This prompt was then run through GPT-4 on the 70 notes and the results were compared to the manually annotated results. All of the results were then compared, and the prompt was updated again based on results from GPT-4. For example, GPT-4 initially misclassified several cases of past housing instability as *current housing instability*. We then updated the prompt to specify that “a patient can only experience a ‘history’ of housing instability if they had housing instability in the past, then were stably housed, then experienced housing instability again. If the note refers to past housing instability, for example, ‘the patient was homeless in the past’, then this can be treated as a ‘history of housing instability’”.

### Model testing and evaluation

Overall, the four methods flagged 25,217 notes from 795 patients from the preliminary positive class as being related to housing and/or housing instability past or present. If a given method flagged more than one note for a specific patient, only the most recent note in relation to the conception date was used for annotation. For example, if GPT-4 flagged two notes dated Jan 1^st^, 2019 and May 1^st,^ 2019, reviewers only annotated the note dated May 1^st^, 2019. This ensures that a maximum of four notes per patient were used for annotation. Model performance was measured by examining accuracy, recall, precision, and F1 score using the Scikit-Learn library.

Because it was initially uncertain how many notes would be flagged by the models for a given number of patients, we initially selected 500 patients from the preliminary positive class. Out of those 500 patients, 295 patients had one or more notes within a 12 month period prior to pregnancy, for a total of 9,451 notes. Of those notes, 511 notes from 139 patients were tagged as either containing information on housing status by the JSL model or information on housing instability past or present by GPT-4, GPT-3.5, and Regex. 204 of the 511 notes were selected as being the most recently note tagged by any of the models for each patient, and all 204 notes were manually annotated. For the second round of model tagging and annotation, 500 patients who had one or more notes within a 12 month period prior to conception were randomly selected from the preliminary positive class. After model tagging, 900 notes from 216 patients were tagged as either containing information on housing status by the JSL model or information on housing instability past or present by GPT-4, GPT-3.5, and Regex. 335 of the 900 notes were selected as being the most recently note tagged by any of the models for each patient, and all 335 notes were manually annotated (positive class). A full breakdown of the number of patients and notes used in each round of model tagging and annotation can be found in Supplementary Table S3.

To identify notes related to housing and/or housing instability past or present from patients in the preliminary negative class, we first selected a random sample of 500 patients from the preliminary negative class. Of those 500 patients, only 348 patients had one or more notes within a 12 month period of their conception date, for a total of 5,455 notes. GPT-4, GPT-3.5, Regex, and JSL were run on all 5,455 notes using the same method as the notes from patients in the positive class. Of the 5,455 notes, a total of 79 notes from 50 patients were flagged by one or more of the four methods. Out of the 79 notes, a total of 59 notes were tagged as being the most recent note tagged by one or more of the models for a given patient. See Supplementary Table S3.

### GPT bias evaluation

FPR (False Positive Rate) and FNR (False Negative Rate) were calculated using FP (false positives), FN (false negatives), TP (true positives), and TN (true negatives), derived from the confusion matrix. FPR: FP/FP+TN. FNR: FN/TP+FN. 95% confidence intervals for a population proportion were calculated using the following formula: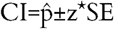, where z= 1.96 for a 95% confidence level. SE (standard error) was calculated using 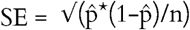, where 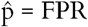 or FNR, and n = sample size.

### De-identification of patient notes

PSJH has an existing corpora of de-identified notes that were created using a sequence of operations performed on text data to remove PHI (protected health information)^34^. These operations included multiple pre-trained ML models and/or regular expressions. Two versions of de-identified notes were used: *complete de-id* in which all PHI was obfuscated and all dates were shifted or masked if shifting was not possible, and *de-id except date*, in which all PHI was obfuscated but the dates were not shifted. All notes remained within the secure PSJH cloud environment.

## Results

### Manual annotation of EHR notes

From the 25,217 notes from the 795 patients, the four automated methods flagged a total of 1,411 notes (Supplementary Table S3). Given how the models were designed, JSL was only able to flag notes related to housing in general (housing noted), while Regex could not distinguish between current and past housing instability. Both GPT-4 and GPT-3.5 were able to flag notes related to general housing status, housing instability current, and housing instability past. After the models were run on the 25,217 notes, we selected notes that were flagged as *housing noted* for JSL, *housing instability current or past* for Regex, and *housing instability current* or *housing instability past* for GPT-4 and GPT-3.5.

The most commonly flagged note types were assessments, plan of care, OB Triage, History and Physical (H&P), consults, discharge summary, and ED (emergency department) notes, indicating that these types of notes are most likely to provide relevant information related to housing and/or housing instability (Supplementary Table S4). We selected the 539 most recent notes from 355 patients for manual annotation. Demographic characteristics of the 355 patients can be found in Supplementary Table S5. Of the 539 manually annotated notes, the most common type of note were progress notes (216) followed by ED provider notes (102) and telephone encounters (38) (Supplementary Table S6).

Of the 182 patients that were identified as having current or past housing instability, only 18% (33 patients) had a structured SNOMED code related to housing instability in their chart (Table 1). Although the percentage of patients with structured codes for HI is low, it is higher than what has been previously reported, and it is well known that structured fields do not adequately capture a patient’s housing status^29,35^. These results could reflect the fact that these notes were selected from patients flagged for HI, or that healthcare teams are more likely to ask about and document housing instability when a patient is pregnant. A total of 10% of patients that were labeled as *stably housed* and 11% of patients labeled *unknown* had a SNOMED code related to housing instability (Table 1). This is likely because researchers only analyzed notes within one year of pregnancy, while the SNOMED code could have been added to a patient’s chart at any time before pregnancy.

**Table 1:** Number of patients in each housing category who either did or did not have a SNOMED code related to housing instability in their chart. SNOMED codes used to identify housing instability can be found in Supplementary Table S1.

|  | SNOMED Code for Homelessness |  |
| --- | --- | --- |
| Housing Label | Yes | No |
| Stably Housed | 11 | 98 |
| Current Housing Instability | 32 | 131 |
| Past Housing Instability | 1 | 18 |
| Unknown | 7 | 57 |

Two annotators manually annotated the 539 notes and flagged 415 as related to housing. Of those 415, 164 were labeled as *stably housed* 223 were labeled as *current housing instability*, 28 were labeled as a *history of housing instability*, and 124 were labeled *unknown* (Table 2). 25% of the notes underwent dual annotation. Before adjudication, dually-annotated notes had a Cohen’s kappa coefficient of 0.589, which reflects moderate agreement^36^, and highlights the ambiguity and subjectivity of annotating complex concepts such as housing instability.

**Table 2:** Percentage of manually annotated notes by housing label.

| Housing Label | Number of Notes | Percentage of Total Notes |
| --- | --- | --- |
| Current Housing Instability | 223 | 41.4 |
| Stably Housed | 164 | 30.4 |
| Unknown | 124 | 23.0 |
| History of Housing Instability | 28 | 5.20 |

### Identification of Current and Past Housing Instability

Because housing status can change over time, it’s important to be able to distinguish between current and past housing instability. This may be especially important for certain groups of patients, such as pregnant women, where housing instability during pregnancy may have different implications than prior housing instability. Figs. 2 and 3 and Supplementary Tables S7 and S8 illustrate the differences in performance metrics between GPT-4, GPT-3.5, RegEx and manual annotation in identifying notes related to current or past housing instability, measured against final adjudicated labels. For current and past housing instability, the recall of GPT-4 was higher (0.924) compared with GPT-3.5 (0.717), RegEx (0.649), and manual annotation (0.702). However, manual annotation had the highest precision amongst the four methods (0.971), compared with GPT-4 (0.850), GPT-3.5 (0.759), and Regex (0.632). The low performance of RegEx was due, in part, to the fact that several acronyms related to housing, such as *SLS* and *RV*, also serve as equivalent medical shorthand for terms such as *single limb support* and *review*. This highlights the shortcomings of using a RegEx-based approach when attempting to identify a complex concept such as housing instability. For identifying current housing instability, GPT-4 still had higher recall than both GPT-3.5 and manual annotation, but both LLMs had lower precision than manual annotation. The recall for GPT-3.5 was higher for current housing instability alone, indicating that this model struggled to identify notes where past housing instability was mentioned, further evidenced in Fig. 5B. This demonstrates that while LLMs have the ability to identify past or current instances of an event such as housing instability, specific models should be tested for their performance in each category individually.

**Fig. 2:**
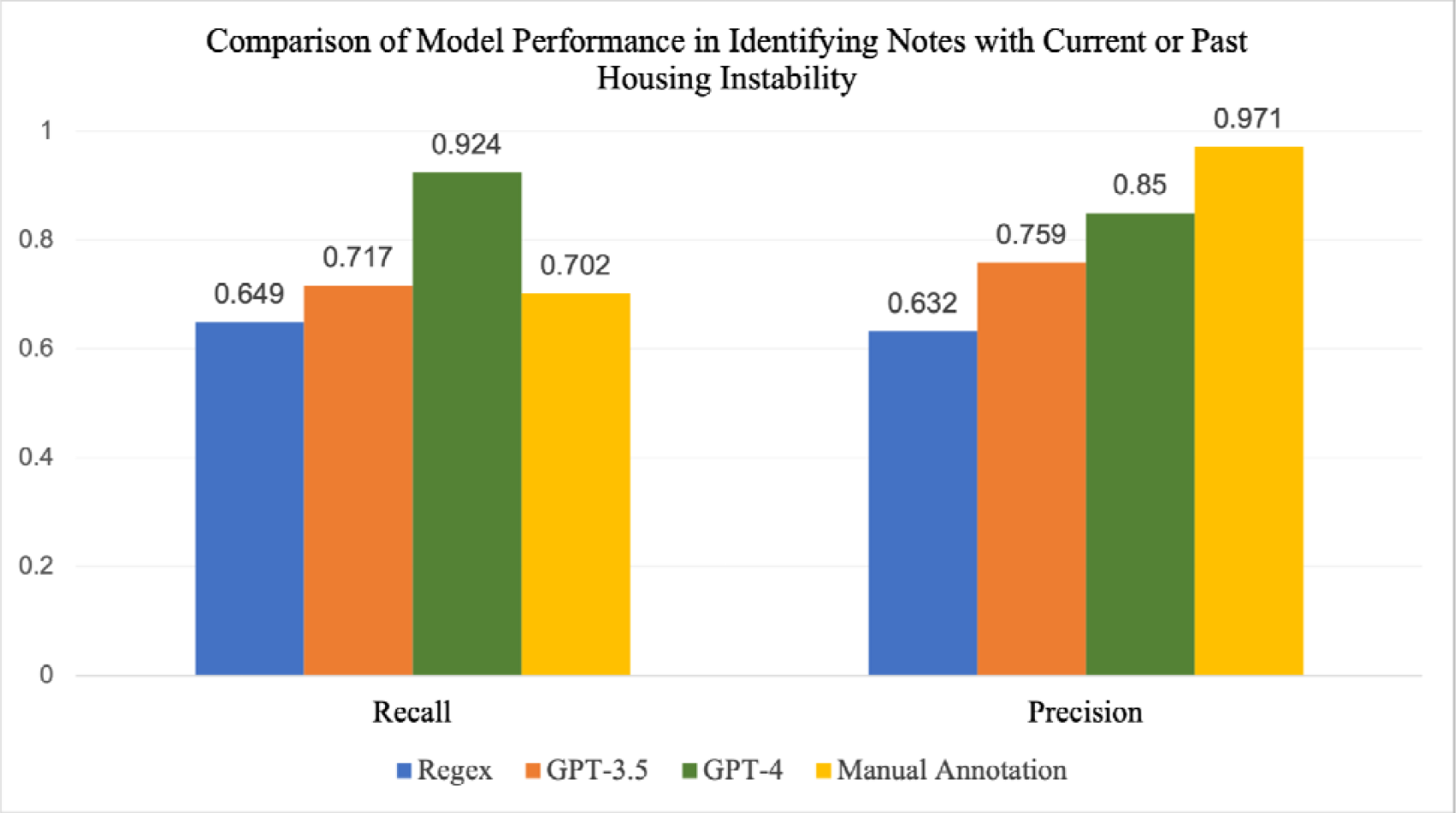
Comparison of recall and precision for Regex, GPT-3.5, GPT-4, and manual annotation in identifying notes with current or past housing instability, measured on 539 manually annotated notes.

**Fig. 3:**
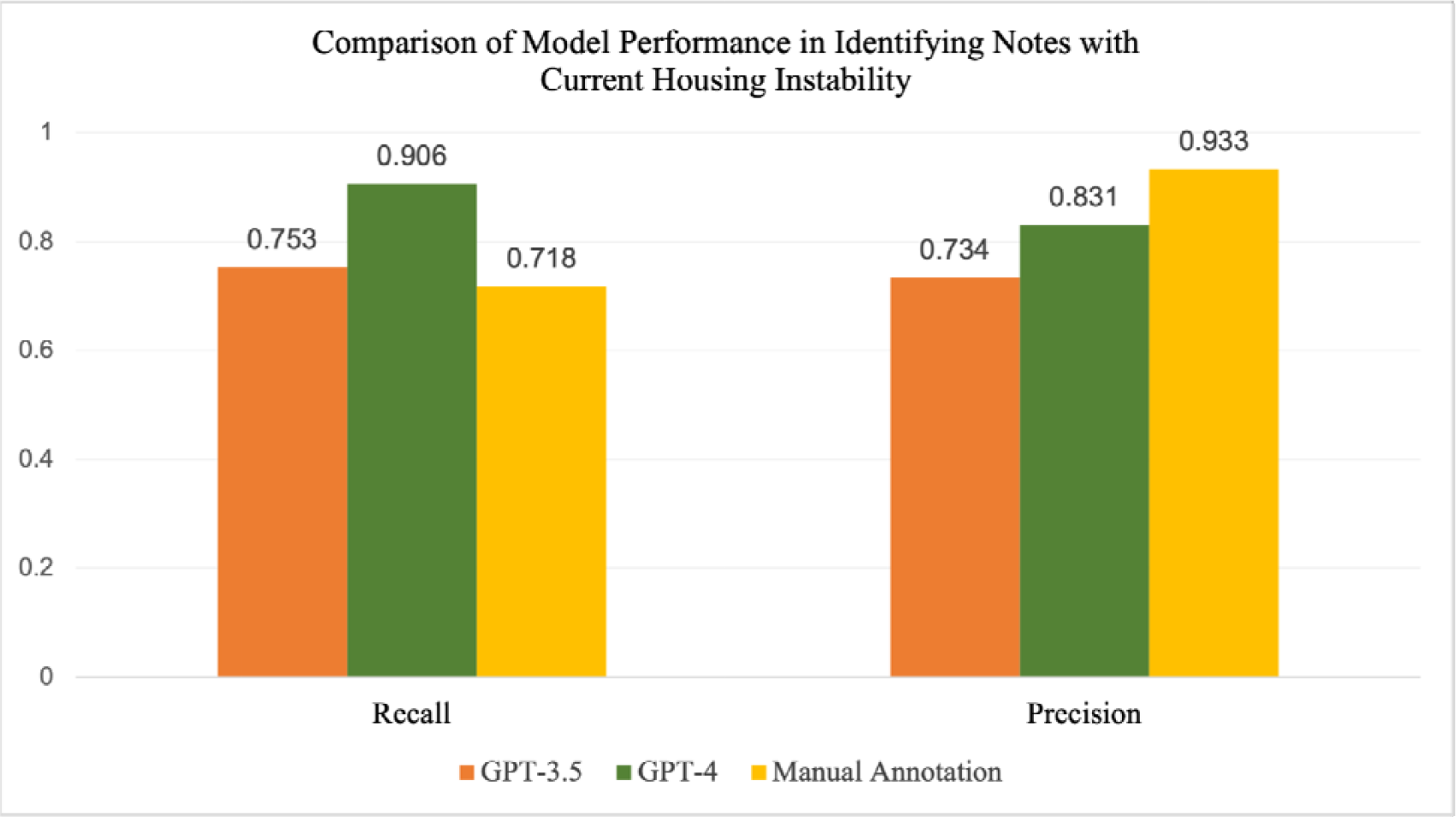
Comparison of recall and precision for GPT-3.5, GPT-4, and manual annotation in identifying notes with current housing instability, measured on 539 manually annotated notes.

The drop in recall for manual annotation highlights the high level of ambiguity associated with annotating this type of information, as well as the energy needed for humans to conduct manual annotation for extended periods of time. Examples of relevant notes, along with the housing labels and justifications from GPT-4 and GPT-3.5 can be found in Table 3. There were several ambiguous cases related to housing in patient notes. For example, “Was in domestic violence. Daughters are [redacted ages] and living with parents for now”. In this example, it is not explicitly stated where the patient is staying, making her current housing status *unknown*. However, this was flagged as *current instability* by GPT-4 and *stably housed* by GPT-3.5. In another example, a patient was living with her family and their landlord was a meth dealer. However, the patient’s boyfriend was actively trying to find them a new living situation, which made the patient happy. While both annotators agreed that the housing situation was not stable, this type of instability was not explicitly mentioned in the note prompt for the LLMs. The reviewers labeled the note as *current instability*, but the note was tagged as *stably housed* by GPT-4 and *current instability* by GPT-3.5.

**Table 3:**
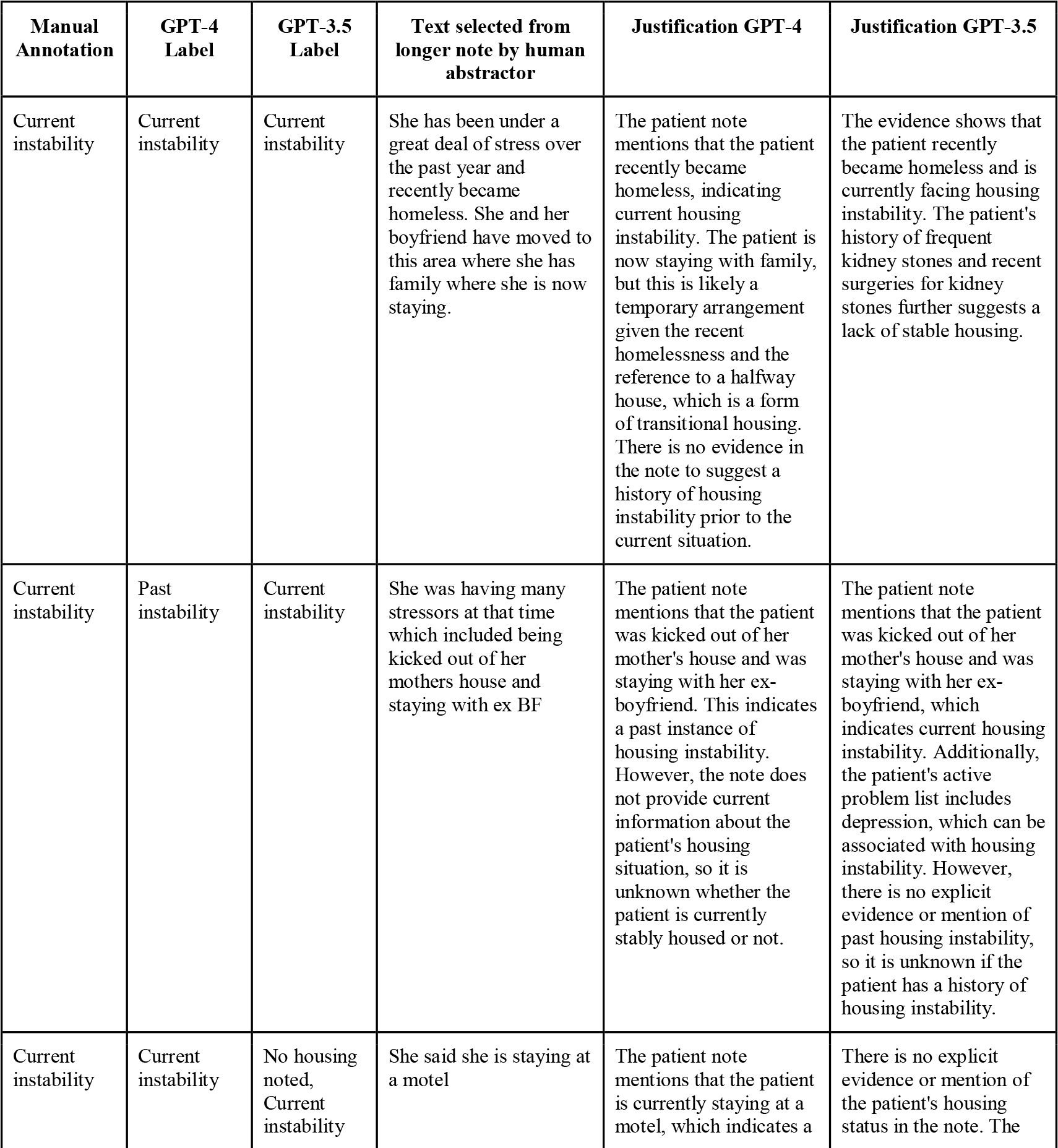
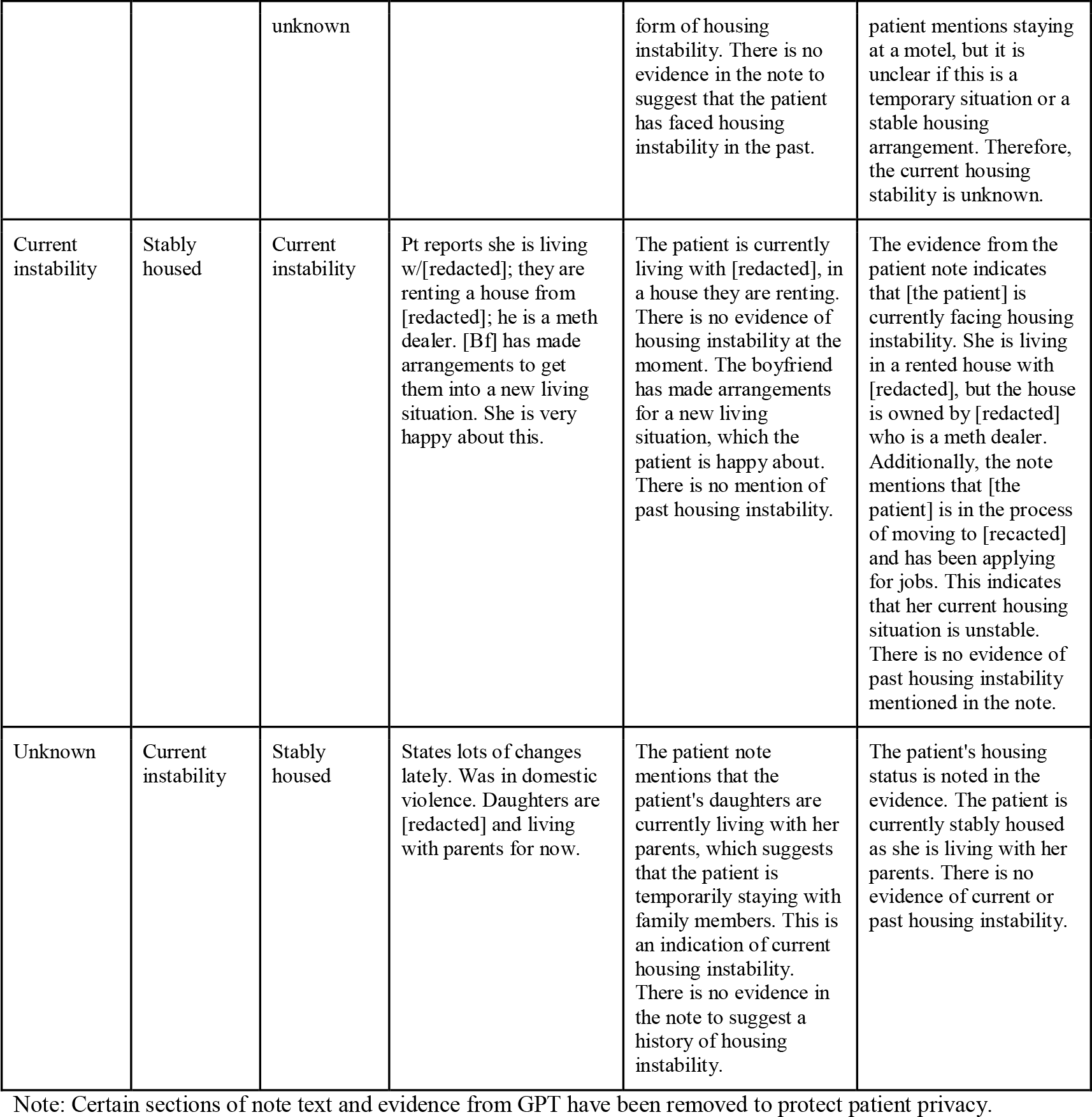
Examples of relevant note text and manual annotation labels compared with GPT-4 and GPT-3.5 and relevant justification.

A minority of notes contained specific references to housing, such as “patient lives in the woods of [redacted location] with her boyfriend”. This phrase signified likely homelessness to most annotators, but one annotator assumed this meant a cabin in the woods. This case was caught by GPT-4 but missed by RegEx and GPT-3.5. There were additional cases where housing instability was explicitly mentioned and was missed by one or more reviewers. This most often occurred in longer notes that contained a significant amount of information, and only 1-2 sentences related to housing, for example “section 8 voucher” which refers to a United States program for assisting very low-income families. This sentence was missed by GPT-3.5 and RegEx but was correctly identified by GPT-4. In addition, there were several notes that contained no information (“blank” notes). In several cases, GPT-3.5 used sentences from the prompt text as evidence and justification, and flagged the note as current or past instability. This did not occur with GPT-4. This is likely because the LLM was asked to provide text evidence verbatim, and GPT-3.5 used the prompt because no relevant note text was available. However, the researchers found no instances of hallucinated evidence in any of the GPT-4 responses that were reviewed, suggesting that requiring verbatim evidence from LLMs can be a solution to hallucinated responses.

Although GPT-3.5 struggled to identify several cases of housing instability, the researchers could not identify a consistent trend in the type or content of the false positive or false negative notes. However, there were several cases where GPT-3.5 listed known risk factors mentioned elsewhere in the note as evidence of housing instability. For example, it noted a patient’s frequent kidney surgeries or a depression diagnosis as justification for HI, although there was clear mention of HI elsewhere in the note (Table 3). This was not the case with GPT-4, which only used direct evidence from the note text that mentioned housing-related terms as justification. Although kidney disease and depression are associated with HI^37–40^, a human annotator would not use this as a justification for HI. Having an LLM use this as justification could be a potential concern, but could also be an opportunity for a different use case, where researchers ask LLMs to identify potential risk factors observed in a set of records. Those results might show bias in LLMs, or highlight patterns that humans overlook. These examples also demonstrate significant differences between the two GPT releases, and highlight the value of providing evidence and justification for every note when using LLMs. In most cases, the evidence gathered by the reviewers was similar or identical, but the interpretation differed. Similarly, the evidence gathered by GPT-4 was similar to the reviewers in most cases (although this was not always the case with GPT-3.5). This indicates that, while manual annotation is likely to yield more accurate results, GPT-4 could be used to rapidly gather relevant note text for computer-assisted manual review, helping save time without losing important or relevant patient information.

### Identification of General Housing Status

Fig. 4 and Supplementary Table S9 shows the performance of GPT-3.5 and GPT-4 compared to JSL and manual annotation in identifying notes where housing was mentioned. GPT-4 outperformed both GPT-3.5 and JSL across all four metrics but had a slightly worse precision compared to manual annotation (0.936 compared with 0.952), although recall was higher (0.781 compared with 0.720). Interestingly, the majority of cases that were missed by GPT-4 were instances where housing was stable, for example, “she lives at home with her children”, or “patient was requesting to go home”. This is likely because the prompt was heavily focused on identifying cases of housing instability, and little guidance was provided on identifying housing status overall. Prompt engineering focused on different proportions of relevant information might yield different and more accurate results.

**Fig. 4:**
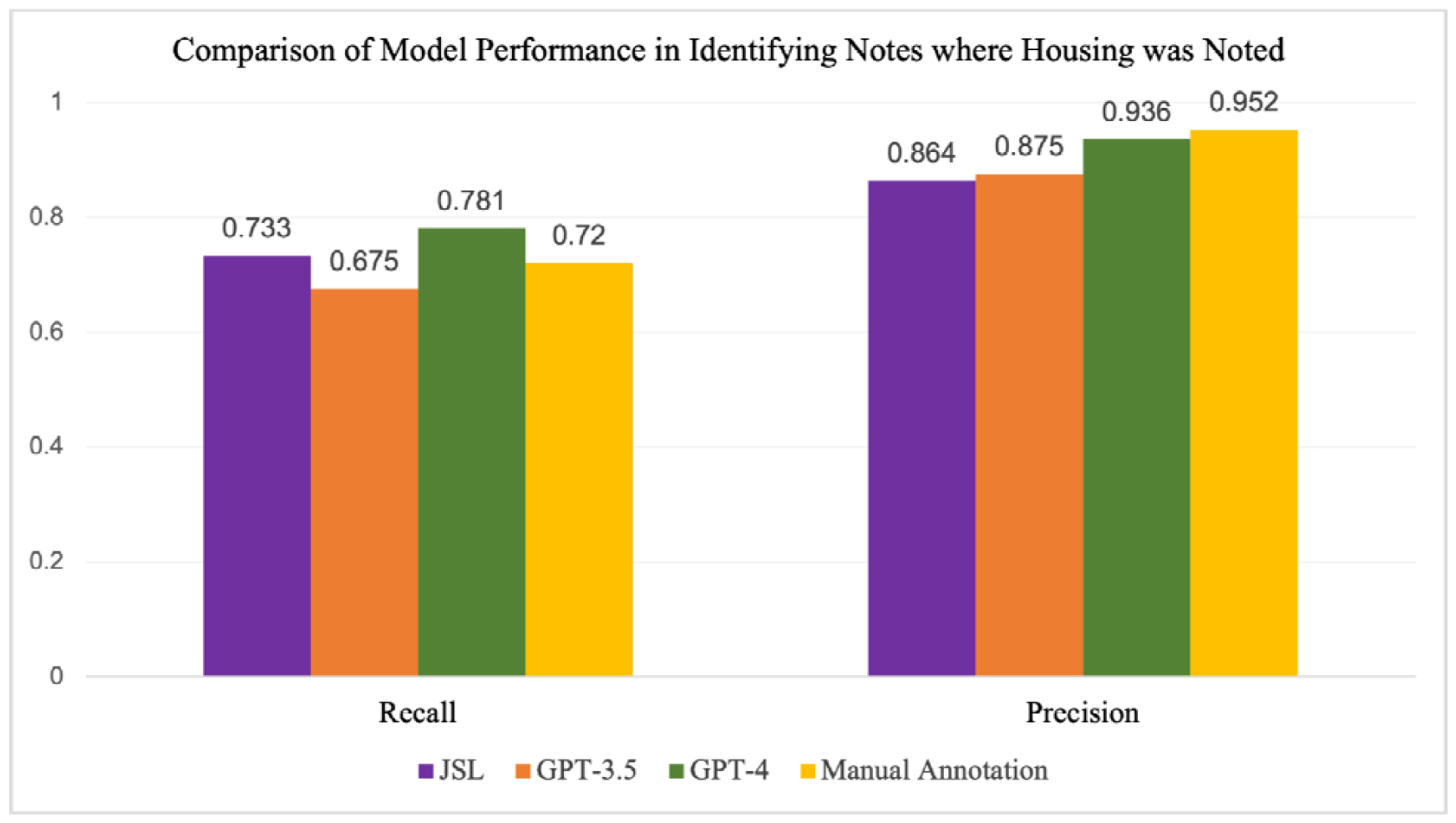
Comparison of recall and precision for JSL, GPT-3.5, GPT-4, and manual annotation in identifying notes where housing was noted, measured on 539 manually annotated notes.

### LLM performance by housing category

Fig. 5 and Supplementary Tables S10 and S11 show the differences in performance for GPT-4 and GPT-3.5 in identifying notes across the different housing categories: stable housing, current housing instability, past housing instability, or unknown. GPT-3.5 performed worse than GPT-4 across all categories and had particularly low recall for notes labeled as *past instability* compared with GPT-4, which had a higher recall than precision in this category. Both GPT-4 and GPT-3.5 demonstrated poor recall for *stable housing* notes; this was likely because the prompt focused more heavily on housing instability compared with stable housing. These data indicate a performance improvement for the GPT-4 release and demonstrate the effects of prompt engineering on the model outcome.

**Fig. 5:**
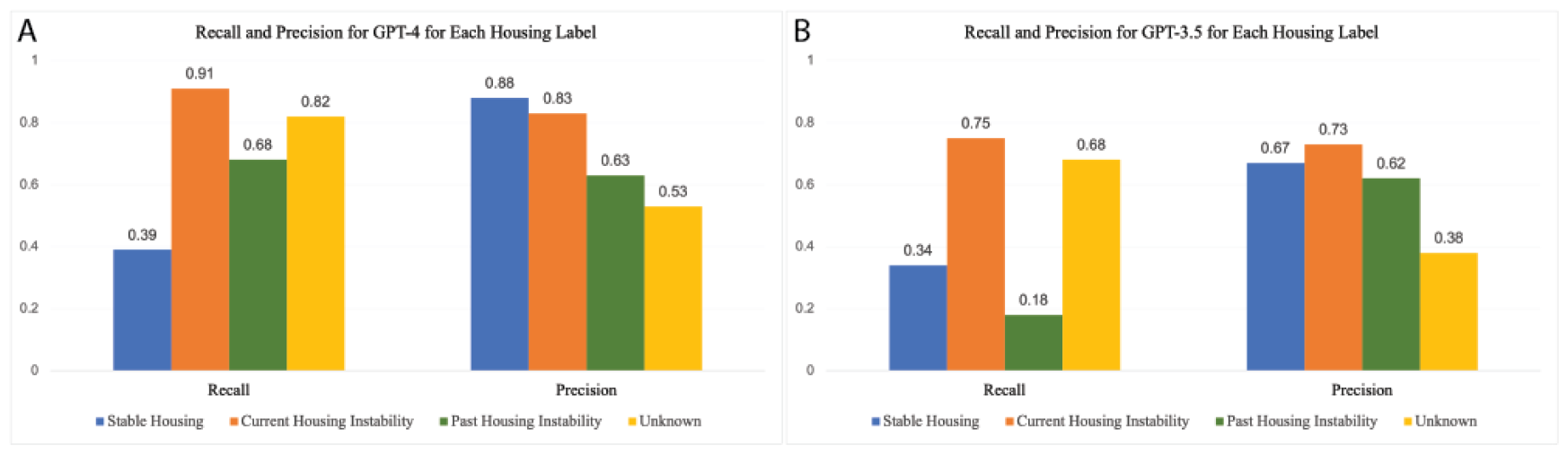
Recall and precision metrics for A) GPT-4 and B) GPT-3.5 for each housing label measured on 539 manually annotated notes.

### LLM bias evaluation

To test for bias in the LLMs, we used the fairness criteria of separation^41^ to examine the false positive rates (FPR) and false negative rates (FNR) between GPT-4 and GPT-3.5 across three different housing labels (housing noted, housing instability past or current, and housing instability current) across the age (18-30 and 31-44), race (White, Black, Asian, AIAN (American Indian & Alaska Native), NHPI (Native Hawaiian and other Pacific Islander), Unknown or Declined, and Other) and ethnic (Hispanic or Latino, not Hispanic or Latino, and Unknown or Declined) demographic groupings. We then examined the 95% confidence intervals of the FPR and FNR for each group (Supplementary Tables S12-S17 and Supplementary Figs. S1-S3). According to the fairness criteria of separation, any difference in FPR and FNR between groups suggests potential algorithmic bias. We did observe differences in FPR and FNR between all of the groups within the three demographic categories. However, when we examined the overlap of the 95% confidence intervals between groups, we found that in all cases, except in cases where the sample sizes were extremely small (n <5), there was overlap between confidence intervals for all the groups, suggesting that the differences between groups are not significant. However, further work with a larger sample population is needed.

### Time and cost breakdown of LLMs compared with manual annotation

To analyze text from the 25,217 notes, GPT-4 took 33 hours and 45 minutes, while GPT-3.5 took 36 hours and 36 minutes. JSL took 40 minutes and Regex took less than one minute. For the 539 manually annotated notes, annotators spent an average of 1.38 minutes per note, taking approximately 12 hours and 39 minutes. This does not include the additional time for adjudication, which varied considerably, from less than 1 minute to more than 20 minutes per note. It is also important to note that many of the clinical notes were very short (1-5 sentences). However, note length can vary, and several notes were multiple paragraphs long. Compared with LLMs, if manual annotators reviewed 25,217 notes, it would have taken approximately 34,800 minutes, or about 580 hours (Fig. 6).

**Fig. 6:**
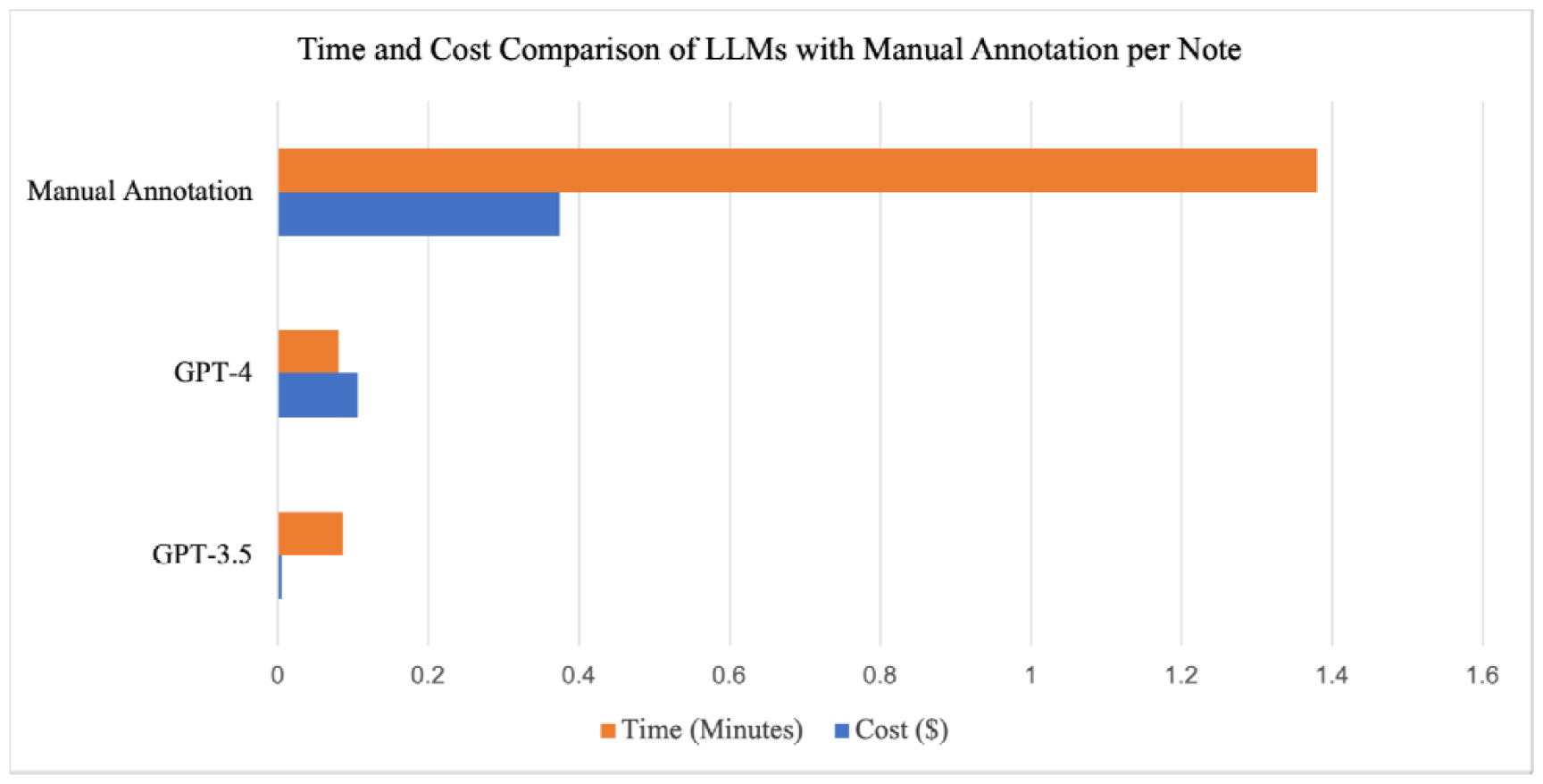
Cost and time comparison of GPT-4 and GPT-3.5 with manual annotation estimate per note..

There was a substantial difference in cost between GPT-4 and GPT-3.5, as shown in Table 4, due to the increase in cost per 1000 tokens for notes for GPT-4 (0.06) compared with GPT-3.5 (0.003). The output cost also increased from 0.004 per 1000 tokens for GPT-3.5 to 0.12 for GPT-4. Interestingly, the prompt (1778.39 USD for GPT-4) cost more than the total for all the notes (701.72 USD for GPT-4), and this was the case for GPT-3.5 as well. This is because the prompt had to be included as part of each note. Because the prompt was long (1,182 tokens), this increased the cost substantially. Future work comparing model performance in relation to prompt length would provide valuable insight into this tradeoff.

**Table 4:**
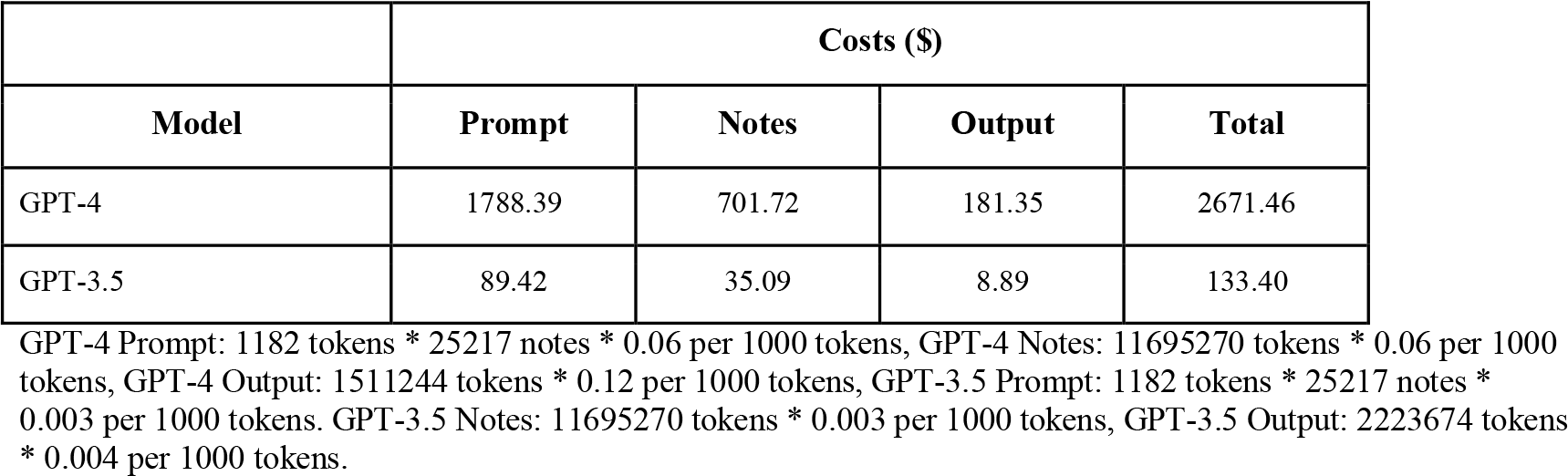
Cost analysis of GPT-4 compared with GPT-3.5 for 25,217 notes from 795 patients.

The cost of manual annotation varies by location, but in the United States can be estimated to be the minimum wage per hour for that state. As of January 2024 in the state of Washington, the minimum wage was $16.28/hr^42^. To analyze 25,217 notes would have cost approximately $9,442, substantially higher than either LLM. An analysis of the time and cost per note between LLMs and manual annotation can be found in Fig. 6.

### Analysis on patients in the preliminary negative class

To evaluate the performance of our four methods compared with patients from the preliminary positive class, we selected a random sample of 5,455 notes from 348 patients in the preliminary negative class. GPT-4, GPT-3.5, Regex, and JSL were run on all 5,455 notes using the same method as notes from patients in the preliminary positive class. Of the 5,455 notes, all four methods only flagged 59 of the most recent notes from 50 patients with one of the four housing labels (GPT-4: 12, GPT-3.5: 11, Regex: 14, JSL: 27).

This demonstrates that, as expected, patients from the preliminary negative class had far fewer notes related to housing and/or housing instability compared with patients in the preliminary positive class. To generate a manually annotated dataset of ∼500 notes to compare with the preliminary positive class, the four methods would need to analyze approximately 46,000 notes. However, JSL is not able to distinguish between notes related to general housing and housing instability, and Regex has low precision and recall for identifying notes related to housing instability. Therefore, the LLMs provide the best chance of finding relevant notes related to housing/housing instability for patients in either class. Because the number of notes flagged by GPT-4 and GPT-3.5 were very low in the preliminary negative class (12 and 11, respectively, out of 5,455 notes), the two models would need to analyze approximately 227,300 notes to find ∼500 notes related to housing and/or housing instability in the preliminary negative class. Due to the cost restrictions of running these LLMs, we were unable to perform this analysis. However, future work to analyze additional notes from patients in the preliminary negative class could provide insight into any differences in notes between these two classes.

### Evaluation of GPT-4 on de-identified patient notes

De-identification can help mitigate privacy risks to individuals to support secondary use of data for research. In the US, the Health Insurance Portability and Accountability Act (HIPAA) specifies 18 categories of information that are protected health information (PHI) that must be removed from medical records^43,44^. However, while the process of de-identification is necessary to protect patient privacy, the information that is removed during this process, such as dates and locations, may result in the loss of important contextual clues needed for LLM analysis of HI. We wanted to examine whether LLMs performed similarly on two versions of de-identified patient notes compared with original notes: 1) fully de-identified notes where all PHI was obfuscated and all dates were shifted or masked if shifting was not possible (hereafter referred to as *complete de-id*), and 2) patient notes where all PHI was obfuscated but the dates were not shifted (hereafter referred to as *de-id no date shift*). All notes remained within the secure PSJH system and no notes were shared publicly. All data processing was conducted in PSJH’s secure cloud environment.

We ran the two de-identified versions of the 539 manually annotated notes through GPT-4 and compared the performance metrics to the original notes to identify current or past housing instability or general housing status (housing noted). We found that in all cases, recall dropped but precision increased for the de-identified notes compared with the original notes (Fig. 7 and Supplementary Table S18). For example, for the notes labeled as *current housing instability*, the recall for GPT-4 on the original notes was 0.906, but this dropped to 0.812 and 0.834 for the complete de-id and de-id date shifted notes, respectively. By contrast, the precision increased from 0.831 for the original notes compared with 0.862 and 0.849 for the complete de-id and de-id date shifted notes, respectively. These minor increases in precision are likely due to the fact that results from GPT-4 are slightly different each time the model is run, resulting in slight differences in performance. The drop in recall is not surprising given the nature of de-identification, in which both places and locations have been obfuscated, making it more difficult for the model to identify relevant notes. For example, there were several cases in the original notes where the patient was stated to be living in a specific location, such as a city or county, but these locations were changed to medical facilities or, in one instance, a jail, resulting in the model sometimes mislabeling the patient as *unstably housed* or missing the note as related to housing altogether. In other cases, when the dates were shifted, instances of *past housing instability* were made current, making it difficult for the LLM to properly identify and label these notes.

**Fig. 7:**
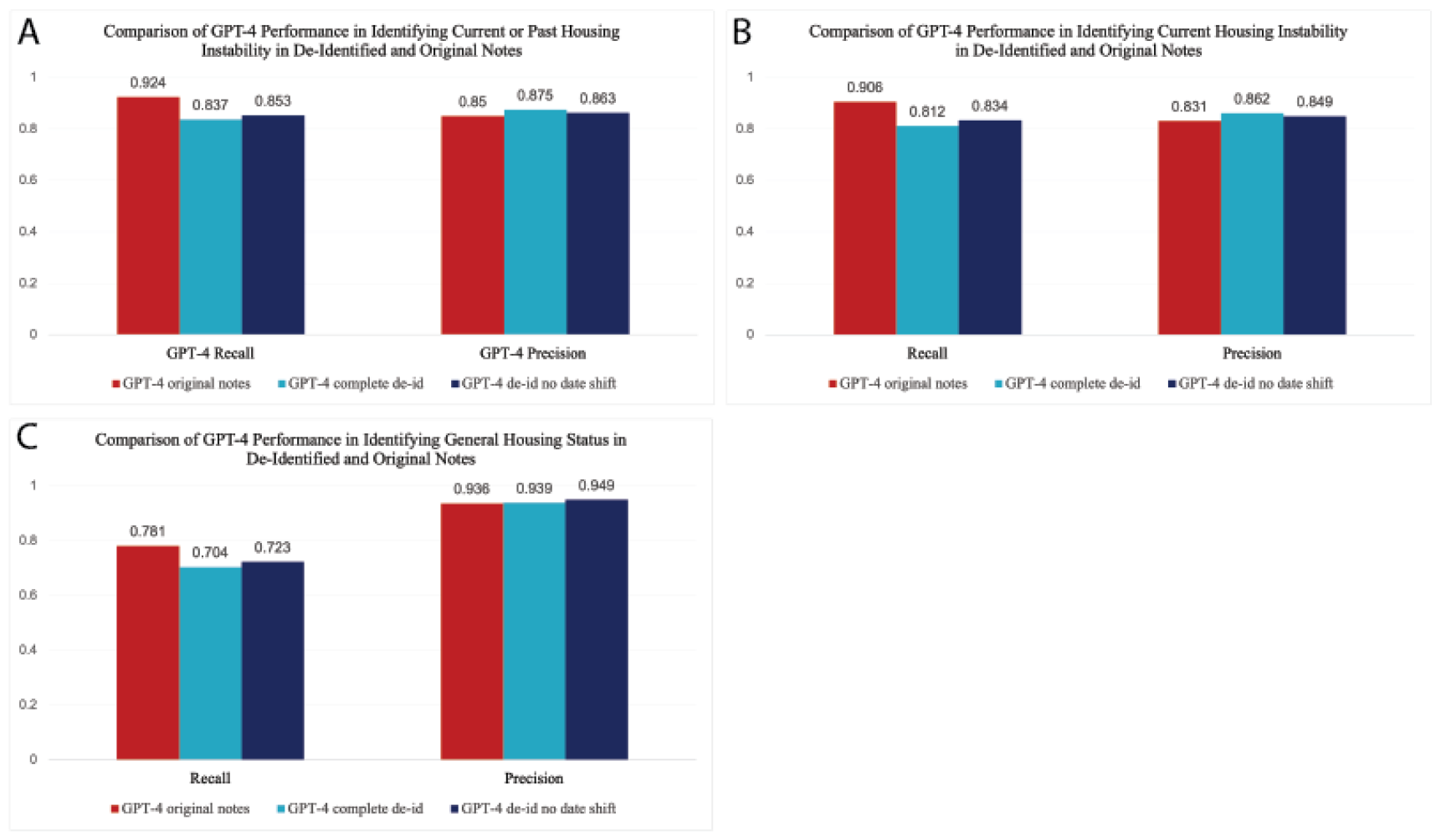
Comparison of GPT-4 performance in identifying A) Current or past housing instability, B) Current housing instability, or C) General housing status from three different versions of the same set of 539 notes. Notes were either complete (original) patient notes, completely de-identified notes, or de-identified notes with no date shift.

## Discussion

Our results demonstrate the value of using LLMs to identify instances of complex SDoH concepts in the EHR, such as past or current housing instability. Although manual annotation correctly classified the most notes related to HI, it took significantly longer and is more expensive than using LLMs, with a minimal increase in performance. GPT-4 outperformed GPT-3.5, JSL and RegEx in identifying patients experiencing current or past housing instability. In most cases, the evidence from GPT-4 was similar or identical to that of the manual annotators, and there was no evidence of hallucinations in GPT-4 output. Our work also suggests that requiring the LLM to provide verbatim evidence and justification from the original text can help to reduce the risk that relevant context about housing information is omitted from LLM results.

It is important to note that housing instability does not exist in a vacuum; oftentimes there are multiple compounding factors that either contribute to, or occur as a result of, housing instability, including domestic violence, drug abuse, and/or mental illness. One limitation of this study was our focus solely on HI and not an additional identification of these risk factors. This resulted in some cases where a patient was technically considered to have stable housing, but there were other risk factors in the patient note that would likely be important for users of abstraction results: case workers, clinicians or researchers. Expanding the prompt might improve performance and enable labeling that separates out multiple dimensions of housing security, including uncertainty about future housing, frequency of housing transitions, and risks from unsafe housing situations. Because GPT-4 and GPT-3.5 are not deterministic models, responses, and therefore performance, may also change if rerun on the same notes. However, the newest release of GPT, GPT-4 Turbo, allows researchers to add a deterministic seed parameter to ensure the model returns the same response every time, helping to prevent changes in performance across multiple runs.

Because all our methods required that each note be analyzed individually, and all four methods identified 1,411 notes out of 25,217 related to housing or HI in the preliminary positive class, and only 64 out of 5,455 notes related to housing or HI in the preliminary negative class, we can conclude that many notes in this study likely did not contain information on housing and/or housing instability. However, future work could investigate the similarities and differences in note content related to housing/HI between patients in both classes. In addition, because we used automated methods for the initial selection of relevant patient notes, we likely missed some patient notes related to housing or HI that were not captured with any of the four automated methods. Future work to manually annotate a larger corpus of patient notes related to housing and HI, as well as other SDoH categories, would prove useful in this regard. Another limitation is that the study was limited to the content documented in EHR notes, and a recent survey reported that only about 60% of patients felt comfortable sharing SDoH-related information^45^. Future studies would benefit from longitudinal confidential surveys or interviews with patients and healthcare teams. In addition, because the time and cost to run GPT-4 and GPT-3.5 might not be feasible across millions of patient notes, work with newly emerging open source language models may provide a similar performance for a much lower cost and runtime.

In conclusion, this work demonstrates that LLMs have potential for computer-assisted annotation of social history, improving recall and reducing costs. This includes temporal feature engineering, such as identifying how long ago a patient may have experienced HI in relation to their pregnancy. Results also identified two important areas where further work is needed: separating out three different dimensions of housing insecurity, and advancing de-identification methods that do not result in loss of social history. Providing greater access to existing SDoH data can be valuable for retrospective research, chart review for prospective trials, and population health interventions to identify those who might benefit from proactive outreach.

## Contributions

AR, NT, MN, and SNP had full access to all data in the study and takes responsibility for the integrity of the data and accuracy of the data analysis. Concept and design: AR, NT, MN, LM, JH. Acquisition, analysis, and interpretation of data: AR, NT, MN, and SNP. Drafting of the manuscript: AR, NT, MN, and SNP. Critical revision of the manuscript: all authors. Statistical analysis: AR and NT. Supervision: JH and LM.

## Data Availability

Results have been aggregated and reported within this paper to the extent possible while maintaining privacy from personal health information as required by US law. All data are archived within Providence St Joseph Health systems in a HIPAA-secure audited compute environment and those wishing to verify study conclusions can contact the Chief Data Officer. All biomedical codes used to extract data from electronic health records have been shared. The code to generate the cohort for the selection of the cohort of pregnant women has been previously described and made publicly available^32^. The code used to run models and reproduce results shown for this study is available at https://github.com/Hadlock-Lab/LLM_SDoH.

## Acknowledgements

Support was provided in part by the National Center for Advancing Translational Sciences, National Institutes of Health, through the Biomedical Data Translator program, award #3OT2TR00344301S1. Any opinions expressed in this document are those of the Translator community at large and do not necessarily reflect the views of NCATS, individual Translator team members, or affiliated organizations and institutions.

