## SupplementaryMaterials for "Using Large Language Models to Annotate Complex Cases of Social Determinants of Health in Longitudinal Clinical Records"

**Appendix: Annotation Guidelines and LLM Prompt	2**

**Supplementary Tables	3**

Supplementary Table S1. SNOMED definitions of conditions.	3

Supplementary Table S2. Regex keywords/phrases for housing instability.	4

Supplementary Table S3. Number of patients and notes used in each round of model tagging and annotation.	4

Supplementary Table S4. Analysis of the different note types of all the notes flagged by GPT-3.5, GPT-4, JSL and/or Regex.	6

Supplementary Table S5. Demographics table of patients with manually annotated notes.	7

Supplementary Table S6. Analysis of the different note types of manually annotated notes.	8

Supplementary Table S7. Model performance metrics in identifying notes that mentioned current or past housing instability.	9

Supplementary Table S8. Model performance metrics in identifying notes that mentioned current housing instability.	9

Supplementary Table S9. Model performance metrics in identifying notes where housing was mentioned.	9

Supplementary Table S10. GPT-4 performance metrics for each housing label.	9

Supplementary Table S11. GPT-3.5 performance metrics for each housing label.	10

Supplementary Table S12. Bias analysis with patient counts for GPT-4 for current and past housing instability.	10

Supplementary Table S13. Bias analysis with patient counts for GPT-3 for current and past housing instability.	11

Supplementary Table S14. Bias analysis with patient counts for GPT-4 for current housing instability.	11

Supplementary Table S15. Bias analysis with patient counts for GPT-3 for current housing instability.	12

Supplementary Table S16. Bias analysis with patient counts for GPT-4 for housing noted.	12

Supplementary Table S17. Bias analysis with patient counts for GPT-3 for housing noted.	13

Supplementary Table S18. GPT-4 performance on original versus de-identified notes for the different housing categories.	13

**Supplementary Figures	15**

Supplementary Figure S1. Bias analysis of GPT-4 and GPT-3.5 for current and past housing instability with confidence intervals.	15

Supplementary Figure S2. Bias analysis of GPT-4 and GPT-3.5 for current housing instability with confidence intervals.	16

Supplementary Figure S3. Bias analysis of GPT-4 and GPT-3.5 for housing noted with confidence intervals.	17

### **Appendix: Annotation Guidelines and LLM Prompt**

**Annotation guidelines for housing stability and housing instability**

People experiencing housing instability are not necessarily experiencing homelessness. Housing instability is often defined to include rent cost burden, risk of eviction, or frequent moves. However, some people who are experiencing housing instability may access homeless services like meal programs, so it is important to distinguish whether an individual spent time in particular homeless service facilities or settings.

Stably housed

- Living in an apartment or home which is paid for by the patient
- Accepted to housing and is preparing to move in
- Permanently living with a family member or friend
  - If no timeline is specified in the note about housing (i.e. not temporary)
  - Examples: Lives with dad. Lives with a friend.
- Patient is discharged to a hospital program with no other mention of housing, example: eating disorder program.

Unknown

- There is no mention of a patient’s housing status
- The information in the note is insufficient to make a final judgment

Unstably housed

- Living in a place not meant for human habitation (the streets, an abandoned building, a vehicle, etc.)
- Recently evicted from their current residence
- Chosen eviction due to unstable home environment
- Physical environment (mold, infestation, etc)
- Living in emergency housing or transitional housing
  - Includes (but not limited to): Group home, foster home
- Temporarily staying with a family member or friend
- Patient’s exact housing status is not explicitly stated, but it is stated that they are facing housing issues or in need of stable housing
- Social work consult for housing
- Patient is worried about future housing insecurity/instability
  - Example: “They’re going to kick me out”

**Final prompt used for GPT-4 and GPT-3.5**

**System Prompt:** You are a social worker reviewing patient notes for social determinants of health. You are looking for patients facing housing instability. Unless the note contains explicit evidence of housing instability, or it can be obviously inferred, you cannot assume a patient is experiencing housing instability.Here is some additional information on homelessness vs housing instability: While patients experiencing homelessness would also be classified as experiencing housing instability, people experiencing housing instability are not necessarily experiencing homelessness. Housing instability is often defined to include rent cost burden, risk of eviction, or frequent moves. Some people who are experiencing housing instability may access homeless services like meal programs, so it is important to distinguish whether an individual spent time in particular homeless service facilities or settings. It is important to note that just because a patient is currently experiencing housing insecurity does not mean that they also experienced it in the past. Unless there are explicit or obviously inferred past references to housing insecurity, or the note is written in a way that implies the patient has been in this situation before, you cannot determine whether or not a patient has a history of housing insecurity. If the note mentions current housing insecurity, for example, 'patient has been homeless for the past two months', this should be treated as 'current' housing insecurity and not 'history'. A patient can only experience a 'history' of housing insecurity if they had housing insecurity in the past, then were stably housed, then experienced housing insecurity again. If the note makes reference to past housing insecurity, for example, 'the patient was homeless in the past', then this can be treated as a 'history of housing insecurity'. **Examples of stable housing: -Living in an apartment or home which is paid for by the patient. -Accepted to housing and is preparing to move in. -Permanently living with a family member or friend. -If no timeline is specified in the note about housing (i.e. not temporary). Examples: Lives with dad, lives with a friend. -Patient is discharged to a hospital program with no other mention of housing. Example: eating disorder program. Examples of unknown: -There is no mention of a patient’s housing status. -The information in the note is insufficient to make a final judgment. Examples of housing insecurity:** -Living in a place not meant for human habitation. Examples: the streets, an abandoned building, a vehicle, etc. -Recently evicted from their current residence. -Chosen eviction due to an unstable home environment. -Chosen or forced eviction due to their physical environment. Examples: mold, infestation, etc. -Living in emergency housing or transitional housing. Examples: Group home, foster home. -Temporarily staying with a family member or friend. -Patient’s exact housing status is not explicitly stated, but it is stated that they are facing housing issues or in need of stable housing. Example: Social work consult for housing. - Patient is worried about future housing insecurity/instability. Example: “They’re going to kick me out”

**Human Message:** Carefully read the following patient note enclosed in triple backticks: **{patient note text}**

Answer the following questions:

The output should be formatted as a JSON instance that conforms to the JSON schema below. As an example, for the schema {"properties": {"foo": {"title": "Foo", "description": "a list of strings", "type": "array", "items": {"type": "string"}}}, "required": ["foo"]} the object {"foo": ["bar", "baz"]} is a well-formatted instance of the schema. The object {"properties": {"foo": ["bar", "baz"]}} is not well-formatted.

Here is the output schema: ``` {"properties": {"Evidence": {"title": "Evidence", "description": "Please provide all evidence of housing status and factors that may be impacting the patient's housing status from the patient note. Please provide evidence verbatim. Include all chunks of text with evidence, not just the first piece of evidence you encounter. Include any information on housing status, whether stable or unstable. Seperate each chunk of text with ' --' and also precede the first chunk of text with '--'. If there is no evidence or housing status is unknown respond by saying \"N/A\". Do not make anything up.", "type": "string"}, "HousingNoted": {"title": "Housingnoted", "description": "Y/N <Is this patient's housing status noted in the evidence?>", "type": "string"}, "HousingInstability_Current": {"title": "Housinginstability Current", "description": "Y/N <Based on the evidence, is this patient currently facing housing instability?. Answer Y/N.>", "type": "string"}, "HousingStability_Current": {"title": "Housingstability Current", "description": "Y/N/Unknown <Based on the evidence, is this patient stably housed? If they are currently facing housing instability then this answer is automatically \"N\". If you do not know, then answer \"Unknown\".>", "type": "string"}, "HousingInstability_History": {"title": "Housinginstability History", "description": "Y/N <Based on the evidence, has this patient faced housing instability in the past, even if their current housing situation is stable?>", "type": "string"}, "Justification": {"title": "Justification", "description": "Justify your responses to the questions above. If there is no evidence or no housing status noted then respond with \"N/A\".", "type": "string"}}, "required": ["Evidence", "HousingNoted", "HousingInstability_Current", "HousingStability_Current", "HousingInstability_History", "Justification"]}

### **Supplementary Tables**

#### Supplementary Table S1. SNOMED definitions of conditions.

| **Condition** | **SNOMED Concept ID** |
| --- | --- |
| Homelessness | 32911000 and all descendent concepts  Homelessness  Finding of temporary shelter arrangements  Sleeping at friends home  Sleeping in night shelter  Sleeping in vehicle  Homeless family  Homeless single person  Housing lack  Lives in squat  Living rough  Sleeping out |

#### Supplementary Table S2. Regex keywords/phrases for housing instability.

homeles[a-z]

houseless

hotel

motel

tent

RVs

encampment

housing insecurity

housing needs

lack of housing

unstabl[a-z] hous[a-z]

SLS

supportive living services

evict[a-z]

couch surfing

shelter

transitional housing

#### Supplementary Table S3. Number of patients and notes used in each round of model tagging and annotation.

|  | Positive Class Annotation Round 1 | Positive Class Annotation Round 2 | All Positive Class Annotation Rounds | Negative Class |
| --- | --- | --- | --- | --- |
| Total Notes Run Through Tagging Models | 9,451 | 15,766 | 25,217 | 5,455 |
| Total Patients Run Through Tagging Models | 295 | 500 | 795 | 348 |
| Notes Tagged by JSL for Housing Status (Most Recent / Total) | 114 / 300 | 169 / 552 | 852 | 33 / 27 |
| Notes Tagged by GPT-3.5 for Housing Instability Past or Present (Most Recent / Total) | 73 / 181 | 115 / 440 | 621 | 12 / 11 |
| Notes Tagged by GPT-4 for Housing Instability Past or Present (Most Recent / Total) | 74 / 284 | 119 / 546 | 830 | 15 / 12 |
| Notes Tagged by Regex Model for Housing Instability Past or Present (Most Recent / Total) | 82 / 226 | 133 / 434 | 660 | 27 / 14 |
| Total Notes that were Tagged by Any Model (Most Recent / Total) | 204 / 511 | 335 / 900 | 539 / 1,411 | 79 / 59 |
| Total Number of Patients with a Flagged Note | 139 | 216 | 355 | 50 |
| Number of Notes that were Manually Annotated | 204 | 335 | 539 | N/A |
| Patients with Notes Selected for Manual Annotation | 139 | 216 | 355 | N/A |

#### **Supplementary Table S4.** Analysis of the different note types of all the notes flagged by GPT-3.5, GPT-4, JSL and/or Regex.

| Note Type | Total number of notes per note type | Number of notes flagged by any model | Percentage of notes flagged by any model |
| --- | --- | --- | --- |
| Progress Notes | 7538 | 660 | 8.76 |
| ED Provider Notes | 1590 | 211 | 13.27 |
| Plan of Care | 1509 | 94 | 6.23 |
| Telephone Encounter | 5951 | 80 | 1.34 |
| ED Notes | 1762 | 80 | 4.54 |
| ED Triage Notes | 1134 | 79 | 6.97 |
| H&P | 306 | 57 | 18.63 |
| Discharge Summary | 222 | 38 | 17.12 |
| Miscellaneous | 418 | 31 | 7.42 |
| Consults | 133 | 24 | 18.05 |
| UC Provider Notes | 177 | 11 | 6.21 |
| OB Triage Notes | 40 | 8 | 20.00 |
| Discharge Instructions | 1080 | 7 | 0.65 |
| Assessment & Plan Note | 517 | 7 | 1.35 |
| Procedures | 152 | 6 | 3.95 |
| Patient Instructions | 871 | 5 | 0.57 |
| L&D Delivery Note | 59 | 3 | 5.08 |
| Anesthesia Preprocedure Evaluation | 128 | 2 | 1.56 |
| Initial Assessments | 4 | 2 | 50.00 |
| Rehab Plan of Care | 2 | 1 | 50.00 |
| Abuse Assessment | 2 | 1 | 50.00 |
| Significant Event | 9 | 1 | 11.11 |
| ED Progress Note | 6 | 1 | 16.67 |
| Treatment Plan | 20 | 1 | 5.00 |
| UC Triage Notes | 177 | 1 | 0.56 |
| Research Note | 1 | 0 | 0.00 |
| Result Encounter Note | 41 | 0 | 0.00 |
| Pulmonary Function | 1 | 0 | 0.00 |
| Sedation Documentation | 3 | 0 | 0.00 |
| Psych | 7 | 0 | 0.00 |
| OR Nursing | 14 | 0 | 0.00 |
| Sleep Disorders | 1 | 0 | 0.00 |
| Subjective & Objective | 1 | 0 | 0.00 |
| UC Notes | 17 | 0 | 0.00 |
| Op Note | 106 | 0 | 0.00 |
| OR PostOp | 4 | 0 | 0.00 |
| ACP (Advance Care Planning) | 1 | 0 | 0.00 |
| Interval H&P Note | 4 | 0 | 0.00 |
| Nursing Note | 4 | 0 | 0.00 |
| Group Note | 3 | 0 | 0.00 |
| Addendum Note | 136 | 0 | 0.00 |
| Anesthesia Pain Management | 4 | 0 | 0.00 |
| Anesthesia Post-op Handoff | 11 | 0 | 0.00 |
| Anesthesia Postprocedure Evaluation | 104 | 0 | 0.00 |
| Anesthesia Procedure Notes | 92 | 0 | 0.00 |
| Brief Op Note | 27 | 0 | 0.00 |
| D-C Instructions Provation | 3 | 0 | 0.00 |
| Goals of Care | 2 | 0 | 0.00 |
| H&P (View-Only) | 6 | 0 | 0.00 |
| Med Student Note | 3 | 0 | 0.00 |
| Historical MyChart Message Note | 9 | 0 | 0.00 |
| Historical Problem List A/P Note | 5 | 0 | 0.00 |
| Hospital Course | 1 | 0 | 0.00 |
| Inpatient Medication Chart | 3 | 0 | 0.00 |
| Interim Summary - Physician | 7 | 0 | 0.00 |
| Interval H&P Note | 20 | 0 | 0.00 |
| Lactation Note | 53 | 0 | 0.00 |
| Legal | 1 | 0 | 0.00 |
| eICU Note | 6 | 0 | 0.00 |

#### **Supplementary Table S5.** Demographics table of patients with manually annotated notes.

| **Demographic** | **Number of patients** | **Percent of total** |
| --- | --- | --- |
| Total | 355 | 100 |
| **Age groups** |  |  |
| 18-30 | 244 | 69 |
| 31-44 | 111 | 31 |
| **Sex** |  |  |
| Female | 355 | 100 |
| **Ethnicity** |  |  |
| Hispanic or Latino | 54 | 15 |
| Not Hispanic or Latino | 296 | 83 |
| Unknown or Declined | 5 | 1 |
| **Race** |  |  |
| AIAN | 16 | 5 |
| Asian | 7 | 2 |
| Black or African American | 18 | 5 |
| NHPI | 12 | 3 |
| White | 263 | 74 |
| Other | 31 | 9 |
| Unknown or declined | 8 | 2 |
| **Residence** |  |  |
| Metropolitan | 304 | 86 |
| Non-metropilotan | 51 | 14 |

#### **Supplementary Table S6.** Analysis of the different note types of manually annotated notes.

| **Note Type** | **Number** |
| --- | --- |
| Progress Notes | 216 |
| ED Provider Notes | 102 |
| Telephone Encounter | 38 |
| ED Triage Notes | 33 |
| ED Notes | 33 |
| Plan of Care | 28 |
| Discharge Summary | 23 |
| H&P | 15 |
| Miscellaneous | 14 |
| Discharge Instructions | 14 |
| UC Provider Notes | 7 |
| Consults | 7 |
| Procedures | 3 |
| OB Triage, Rehab Plan of Care, Assessment & Plan, Initial Assessments, UC Triage, Treatment Plan, Patient Instructions | 1 |

#### Supplementary Table S7. Model performance metrics in identifying notes that mentioned current or past housing instability.

| **Method** | **Accuracy** | **Recall** | **Precision** | **F1 Score** |
| --- | --- | --- | --- | --- |
| Regex | 0.660 | 0.649 | 0.632 | 0.640 |
| GPT-3.5 | 0.763 | 0.717 | 0.759 | 0.738 |
| GPT-4 | 0.889 | 0.924 | 0.850 | 0.885 |
| Manual Annotation | 0.861 | 0.702 | 0.971 | 0.815 |

Regex, GPT-3.5, GPT-4: Positive class = 251, Negative class = 288. Manual annotation: Positive class = 47, Negative class = 61.

#### **Supplementary Table S8.** Model performance metrics in identifying notes that mentioned current housing instability.

| **Method** | **Accuracy** | **Recall** | **Precision** | **F1 Score** |
| --- | --- | --- | --- | --- |
| GPT-3.5 | 0.785 | 0.753 | 0.734 | 0.743 |
| GPT-4 | 0.885 | 0.906 | 0.831 | 0.867 |
| Manual Annotation | 0.880 | 0.718 | 0.933 | 0.812 |

GPT-3.5, GPT-4: Positive class = 223, Negative class = 316. Manual annotation: Positive class = 39, Negative class = 69.

#### **Supplementary Table S9.** Model performance metrics in identifying notes where housing was mentioned.

| **Method** | **Accuracy** | **Recall** | **Precision** | **F1 Score** |
| --- | --- | --- | --- | --- |
| JSL | 0.705 | 0.733 | 0.864 | 0.793 |
| GPT-3.5 | 0.675 | 0.675 | 0.875 | 0.762 |
| GPT-4 | 0.790 | 0.781 | 0.936 | 0.852 |
| Manual Annotation | 0.759 | 0.720 | 0.952 | 0.819 |

JSL, GPT-3.5, GPT-4: Positive class = 415, Negative class = 124. Manual annotation: Positive class = 82, Negative class = 26.

#### Supplementary Table S10. GPT-4 performance metrics for each housing label.

| **Label** | **Recall** | **Precision** | **F1 Score** | **Support** |
| --- | --- | --- | --- | --- |
| Stable Housing | 0.390 | 0.877 | 0.540 | 164 |
| Current Housing Instability | 0.906 | 0.831 | 0.867 | 223 |
| Past Housing Instability | 0.679 | 0.633 | 0.655 | 28 |
| Unknown | 0.823 | 0.528 | 0.644 | 124 |

#### **Supplementary Table S11.** GPT-3.5 performance metrics for each housing label.

| **Label** | **Recall** | **Precision** | **F1 Score** | **Support** |
| --- | --- | --- | --- | --- |
| Stable Housing | 0.341 | 0.675 | 0.453 | 164 |
| Current Housing Instability | 0.753 | 0.734 | 0.743 | 223 |
| Past Housing Instability | 0.179 | 0.625 | 0.278 | 28 |
| Unknown | 0.677 | 0.384 | 0.490 | 124 |

#### **Supplementary Table S12.** Bias analysis with patient counts for GPT-4 for current and past housing instability.

| **Demographic Category** | **Group** | **FPR GPT-4** | **FNR GPT-4** | **Count: True Negative** | **Count: True Positive** | **Count: False Negative** | **Count: False Positive** |
| --- | --- | --- | --- | --- | --- | --- | --- |
| Age | 18-30 | 0.148 | 0.066 | 167 | 169 | 12 | 29 |
|  | 31-44 | 0.130 | 0.100 | 80 | 63 | 7 | 12 |
| Ethnic Group | Not Hispanic or Latino | 0.141 | 0.074 | 207 | 200 | 16 | 34 |
|  | Hispanic or Latino | 0.091 | 0.094 | 40 | 29 | 3 | 4 |
|  | Unknown or Declined | 1 | 0 | 0 | 3 | 0 | 3 |
| Race | White | 0.155 | 0.073 | 180 | 165 | 13 | 33 |
|  | Other | 0.077 | 0.045 | 24 | 21 | 1 | 2 |
|  | Black | 0.154 | 0.188 | 11 | 13 | 3 | 2 |
|  | AIAN | 0.091 | 0.071 | 10 | 13 | 1 | 1 |
|  | NHPI | 0.250 | 0 | 9 | 11 | 0 | 3 |
|  | Unknown or Declined | 0 | 0.111 | 3 | 8 | 1 | 0 |
|  | Asian | 0 | 0 | 10 | 1 | 0 | 0 |

#### Supplementary Table S13. Bias analysis with patient counts for GPT-3 for current and past housing instability.

| **Demographic Category** | **Group** | **FPR GPT-3.5** | **FNR GPT-3.5** | **Count: True Negative** | **Count: True Positive** | **Count: False Negative** | **Count: False Positive** |
| --- | --- | --- | --- | --- | --- | --- | --- |
| Age | 18-30 | 0.179 | 0.298 | 161 | 127 | 54 | 35 |
|  | 31-44 | 0.239 | 0.243 | 70 | 53 | 17 | 22 |
| Ethnic Group | Not Hispanic or Latino | 0.212 | 0.273 | 190 | 157 | 59 | 51 |
|  | Hispanic or Latino | 0.114 | 0.375 | 39 | 20 | 12 | 5 |
|  | Unknown or Declined | 0.333 | 0 | 2 | 3 | 0 | 1 |
| Race | White | 0.207 | 0.258 | 169 | 132 | 46 | 44 |
|  | Other | 0.038 | 0.364 | 25 | 14 | 8 | 1 |
|  | Black | 0.231 | 0.375 | 10 | 10 | 6 | 3 |
|  | AIAN | 0.273 | 0.286 | 8 | 10 | 4 | 3 |
|  | NHPI | 0.167 | 0.364 | 10 | 7 | 4 | 2 |
|  | Unknown or Declined | 0.667 | 0.333 | 1 | 6 | 3 | 2 |
|  | Asian | 0.200 | 0 | 8 | 1 | 0 | 2 |

#### **Supplementary Table S14.** Bias analysis with patient counts for GPT-4 for current housing instability.

| **Demographic Category** | **Group** | **FPR GPT-4** | **FNR GPT-4** | **Count: True Negative** | **Count: True Positive** | **Count: False Negative** | **Count: False Positive** |
| --- | --- | --- | --- | --- | --- | --- | --- |
| Age | 18-30 | 0.135 | 0.093 | 186 | 147 | 15 | 29 |
|  | 31-44 | 0.119 | 0.098 | 89 | 55 | 6 | 12 |
| Ethnic Group | Not Hispanic or Latino | 0.130 | 0.092 | 227 | 178 | 18 | 34 |
|  | Hispanic or Latino | 0.080 | 0.115 | 46 | 23 | 3 | 4 |
|  | Unknown or Declined | 0.600 | 0 | 2 | 1 | 0 | 3 |
| Race | White | 0.148 | 0.105 | 195 | 145 | 17 | 34 |
|  | Other | 0.065 | 0.059 | 29 | 16 | 1 | 2 |
|  | Black | 0.062 | 0.154 | 15 | 11 | 2 | 1 |
|  | AIAN | 0.091 | 0.071 | 10 | 13 | 1 | 1 |
|  | NHPI | 0.231 | 0 | 10 | 10 | 0 | 3 |
|  | Unknown or Declined | 0 | 0 | 6 | 6 | 0 | 0 |
|  | Asian | 0 | 0 | 10 | 1 | 0 | 0 |

#### **Supplementary Table S15.** Bias analysis with patient counts for GPT-3 for current housing instability.

| **Demographic Category** | **Group** | **FPR GPT-3.5** | **FNR GPT-3.5** | **Count: True Negative** | **Count: True Positive** | **Count: False Negative** | **Count: False Positive** |
| --- | --- | --- | --- | --- | --- | --- | --- |
| Age | 18-30 | 0.167 | 0.265 | 179 | 119 | 43 | 36 |
|  | 31-44 | 0.248 | 0.197 | 76 | 49 | 12 | 25 |
| Ethnic Group | Not Hispanic or Latino | 0.207 | 0.245 | 207 | 148 | 48 | 54 |
|  | Hispanic or Latino | 0.120 | 0.269 | 44 | 19 | 7 | 6 |
|  | Unknown or Declined | 0.200 | 0 | 4 | 1 | 0 | 1 |
| Race | White | 0.205 | 0.235 | 182 | 124 | 38 | 47 |
|  | Other | 0.065 | 0.294 | 29 | 12 | 5 | 2 |
|  | Black | 0.188 | 0.231 | 13 | 10 | 3 | 3 |
|  | AIAN | 0.273 | 0.286 | 8 | 10 | 4 | 3 |
|  | NHPI | 0.154 | 0.300 | 11 | 7 | 3 | 2 |
|  | Unknown or Declined | 0.333 | 0.333 | 4 | 4 | 2 | 2 |
|  | Asian | 0.200 | 0 | 8 | 1 | 0 | 2 |

#### **Supplementary Table S16**. Bias analysis with patient counts for GPT-4 for housing noted.

| **Demographic Category** | **Group** | **FPR GPT-4** | **FNR GPT-4** | **Count: True Negative** | **Count: True Positive** | **Count: False Negative** | **Count: False Positive** |
| --- | --- | --- | --- | --- | --- | --- | --- |
| Age | 18-30 | 0.173 | 0.220 | 67 | 231 | 65 | 14 |
|  | 31-44 | 0.186 | 0.218 | 35 | 93 | 26 | 8 |
| Ethnic Group | Not Hispanic or Latino | 0.154 | 0.204 | 88 | 281 | 72 | 16 |
|  | Hispanic or Latino | 0.176 | 0.322 | 14 | 40 | 19 | 3 |
|  | Unknown or Declined | 1 | 0 | 0 | 3 | 0 | 3 |
| Race | White | 0.207 | 0.217 | 69 | 238 | 66 | 18 |
|  | Other | 0.001 | 0.237 | 9 | 29 | 9 | 1 |
|  | Black | 0 | 0.208 | 5 | 19 | 5 | 0 |
|  | AIAN | 0.125 | 0.176 | 7 | 14 | 3 | 1 |
|  | NHPI | 0.333 | 0.235 | 4 | 13 | 4 | 2 |
|  | Unknown or Declined | 0 | 0.100 | 2 | 9 | 1 | 0 |
|  | Asian | 0 | 0.600 | 6 | 2 | 3 | 0 |

#### **Supplementary Table S17.** Bias analysis with patient counts for GPT-3 for housing noted.

| **Demographic Category** | **Group** | **FPR GPT-3.5** | **FNR GPT-3.5** | **Count: True Negative** | **Count: True Positive** | **Count: False Negative** | **Count: False Positive** |
| --- | --- | --- | --- | --- | --- | --- | --- |
| Age | 18-30 | 0.284 | 0.331 | 58 | 198 | 98 | 23 |
|  | 31-44 | 0.395 | 0.311 | 26 | 82 | 37 | 17 |
| Ethnic Group | Not Hispanic or Latino | 0.327 | 0.320 | 70 | 240 | 113 | 34 |
|  | Hispanic or Latino | 0.294 | 0.373 | 12 | 37 | 22 | 5 |
|  | Unknown or Declined | 0.333 | 0 | 2 | 3 | 0 | 1 |
| Race | White | 0.322 | 0.316 | 59 | 208 | 96 | 28 |
|  | Other | 0.100 | 0.316 | 9 | 26 | 12 | 1 |
|  | Black | 0.400 | 0.375 | 3 | 15 | 9 | 2 |
|  | AIAN | 0.375 | 0.235 | 5 | 13 | 4 | 3 |
|  | NHPI | 0.167 | 0.471 | 5 | 9 | 8 | 1 |
|  | Unknown or Declined | 1 | 0.200 | 0 | 8 | 2 | 2 |
|  | Asian | 0.500 | 0.800 | 3 | 1 | 4 | 3 |

#### **Supplementary Table S18.** GPT-4 performance on original versus de-identified notes for the different housing categories.

| **Housing Instability Current or Past** | | |
| --- | --- | --- |
| Notes | Recall | Precision |
| GPT-4 original notes | 0.924 | 0.850 |
| GPT-4 complete de-id | 0.837 | 0.875 |
| GPT-4 de-id no date shift | 0.853 | 0.863 |
| **Housing Instability Current** | | |
| GPT-4 original notes | 0.906 | 0.831 |
| GPT-4 complete de-id | 0.812 | 0.862 |
| GPT-4 de-id no date shift | 0.834 | 0.849 |
| **Housing Noted** | | |
| GPT-4 original notes | 0.781 | 0.936 |
| GPT-4 complete de-id | 0.704 | 0.939 |
| GPT-4 de-id no date shift | 0.723 | 0.949 |

### **Supplementary Figures**

#### Supplementary Figure S1. Bias analysis of GPT-4 and GPT-3.5 for current and past housing instability with confidence intervals.

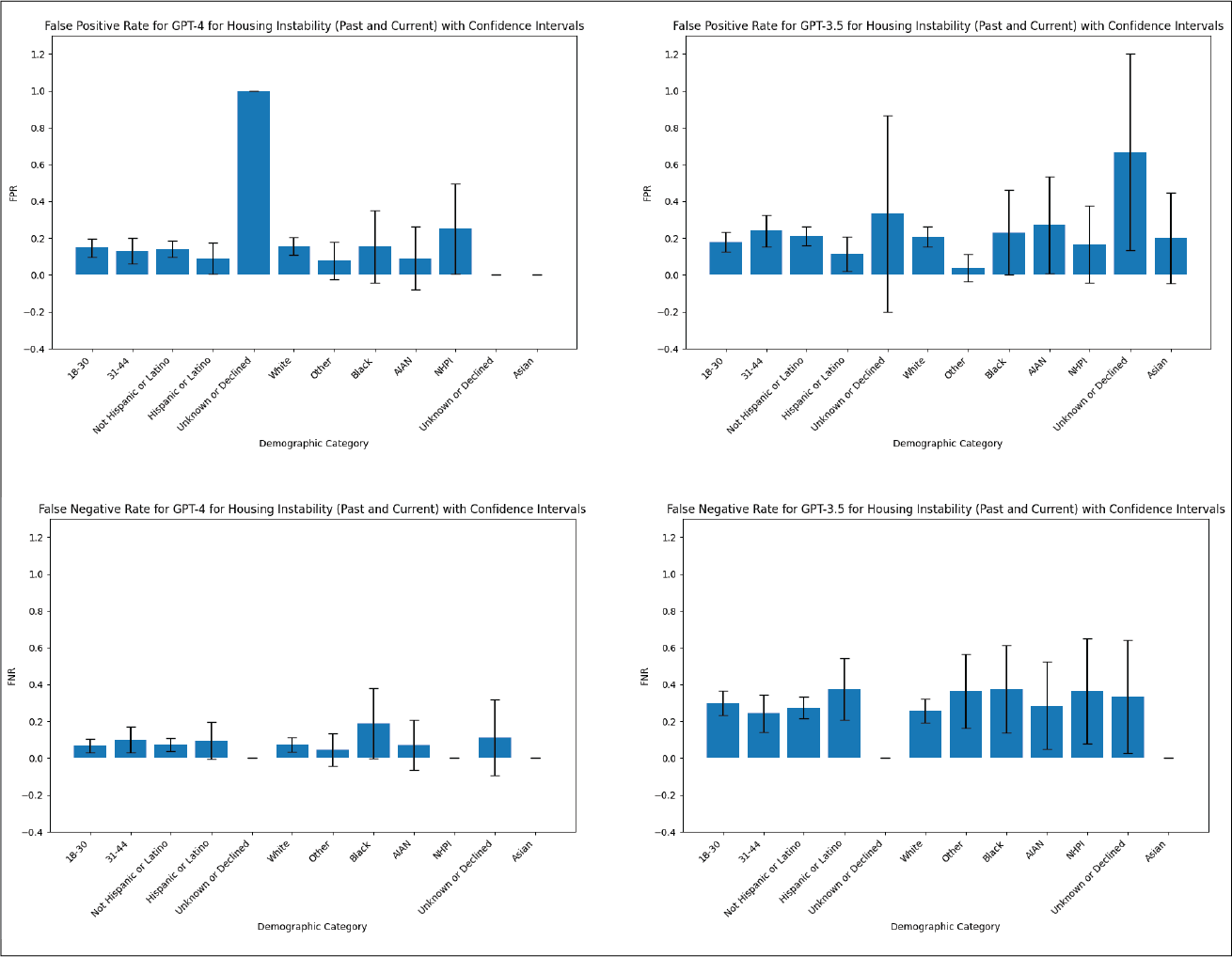

AIAN: American Indian & Alaska Native, NHPI: Native Hawaiian and other Pacific Islander. The first category of “Unknown or Declined” refers to ethnicity, the second category of “Unknown or Declined” refers to race.

#### Supplementary Figure S2. Bias analysis of GPT-4 and GPT-3.5 for current housing instability with confidence intervals.

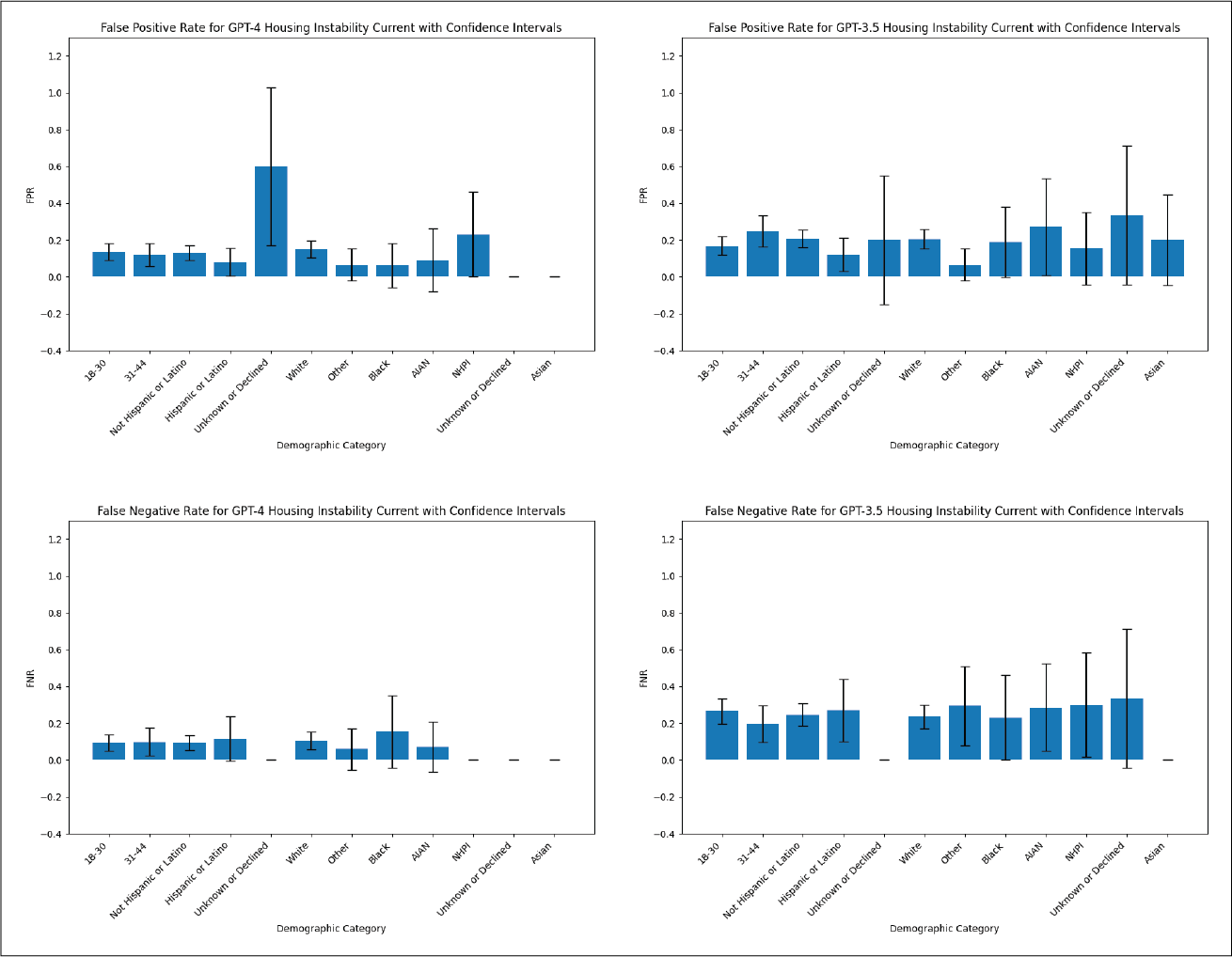

AIAN: American Indian & Alaska Native, NHPI: Native Hawaiian and other Pacific Islander. The first category of “Unknown or Declined” refers to ethnicity, the second category of “Unknown or Declined” refers to race.

#### Supplementary Figure S3. Bias analysis of GPT-4 and GPT-3.5 for housing noted with confidence intervals.

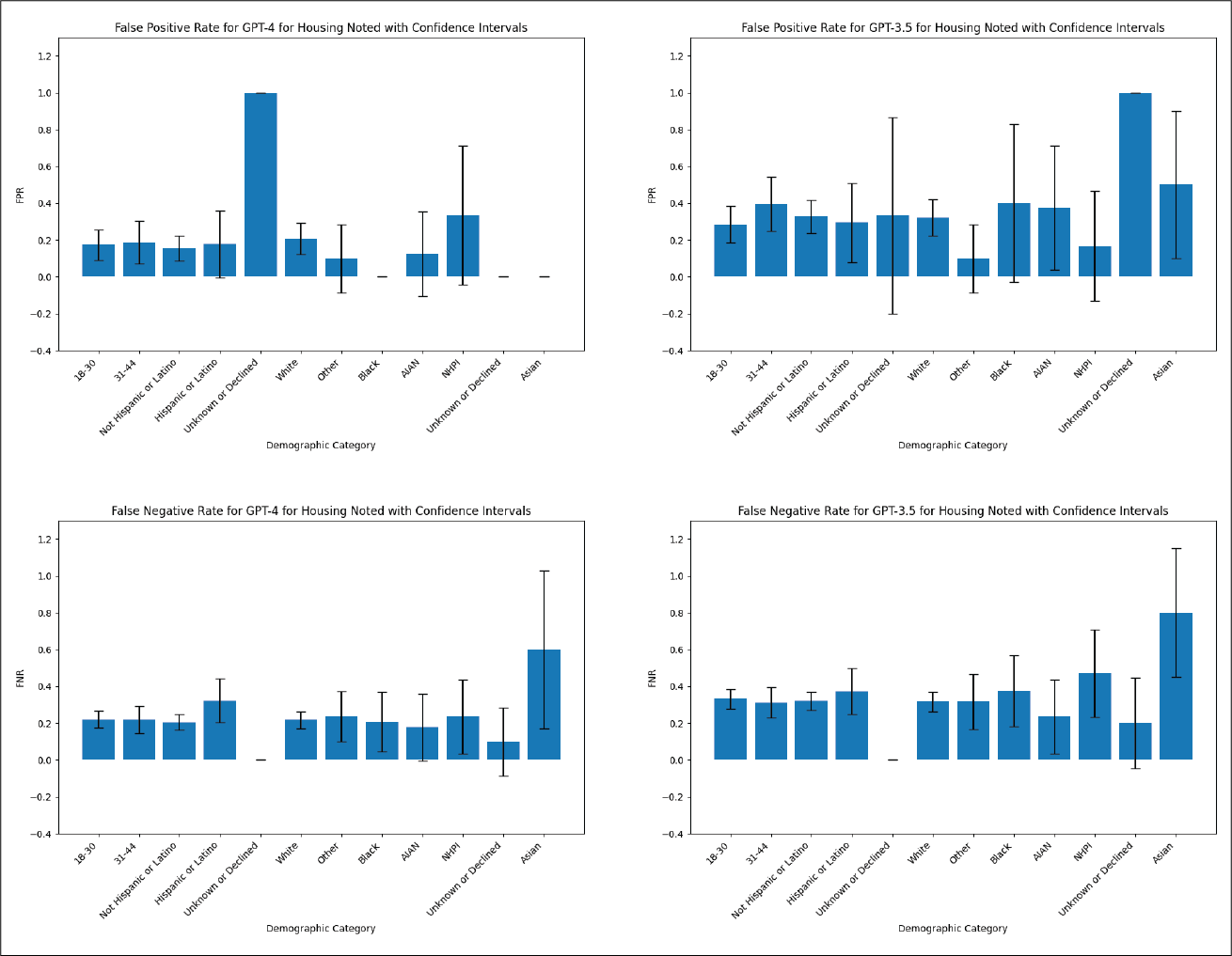

AIAN: American Indian & Alaska Native, NHPI: Native Hawaiian and other Pacific Islander. The first category of “Unknown or Declined” refers to ethnicity, the second category of “Unknown or Declined” refers to race.
